# Population variation and prognostic potential of gut antibiotic resistome

**DOI:** 10.1101/2024.08.08.24311663

**Authors:** K. Pärnänen, M. Ruuskanen, G. Sommeria-Klein, V. Laitinen, P. Kantanen, G. Méric, C. Gazolla Volpiano, M. Inouye, R. Knight, V. Salomaa, A. S. Havulinna, T. Niiranen, L. Lahti

## Abstract

The spread of antibiotic-resistance genes in bacteria has severely reduced the efficacy of antibiotics, now contributing to 1.3 million deaths annually. Despite the far-reaching epidemiological implications of this trend, the extent to which antimicrobial resistance load varies within human populations and the drivers that contribute most to this variation remain unclear. Here, we demonstrate in a representative cohort of 7,095 Finnish adults^1^ that socio-demographic factors, lifestyle, and gut microbial community composition shape resistance selection and transmission processes. Antimicrobial resistance gene load was linked not only to prior use of antibiotics, as anticipated, but also to frequent consumption of fresh vegetables and poultry, two food groups previously reported to contain antibiotic-resistant bacteria. Interestingly, ARG load was not associated with high-fat and -sugar foods. Furthermore, antimicrobial resistance gene load was systematically higher in females and the generally healthier high-income demographics in urban and densely populated areas. Data from this prospective cohort with a 17-year follow-up suggests that the prognostic potential of antimicrobial resistome is comparable to blood pressure for mortality and sepsis. These findings highlight population-level risks and socio-demographic dimensions of antimicrobial resistance that are particularly relevant in the current context of global urbanization and middle-class growth.

## INTRODUCTION

Antibiotic-resistant bacteria (ARB) pose a rapidly increasing threat to global health due to failures in treating infections. By 2050, antimicrobial resistance (AMR, including antibiotic, antifungal, antiparasitic, and antiviral resistance) is predicted to contribute to 10 million deaths annually, surpassing the current most common causes of death, such as cancer and cardiovascular disease^2^. The resistance is encoded by multiple classes of antibiotic resistance genes (ARGs) that can spread across bacterial and human populations. In particular, the human gut has been established as a major reservoir of ARGs and a highway for their lateral transfer^3,4^. The entire collection of ARGs in a given habitat, such as the gut, is called the *resistome*^5,6^.

The emergence and spread of antibiotic resistance in microbial communities^3,7,8^ are driven by the ecological processes of *selection* and *transmission* (Figure 1), which act on both the resistance genes and the bacteria that carry them. *Direct selection* of genes that confer antibiotic resistance can be caused, for instance, by antibiotic use, as it favors the selection of resistant strains and, thus, ARG proliferation in the human gut resistome. *Indirect selection* of resistant bacteria can be caused by environmental factors that influence the growth of bacterial communities with specific types of resistomes^9^. Finally, *transmission* of resistance genes or resistant bacteria between individuals has been suggested to be a key driver of clinical resistance^10^. In particular, previous studies have indicated that contaminated food, person-to-person contact, and travel are possible risk factors for acquiring ARGs^11–13^.

**Figure 1:**
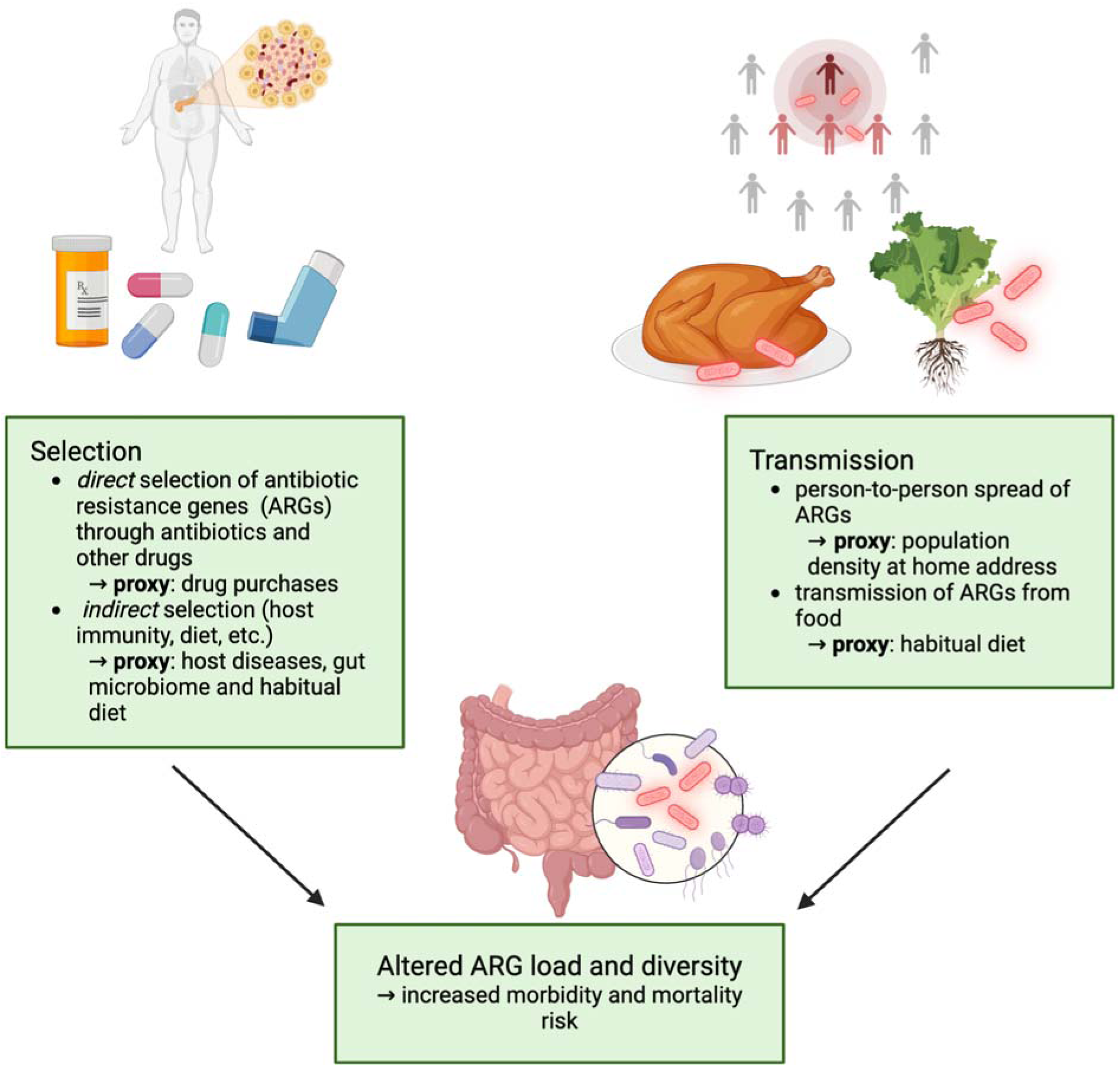
Ecological and epidemiological framework for population-level variation in ARG load and resistome. The ecological processes underlying ARG variation include 1) *selection* of antimicrobial resistance genes (ARGs) and bacteria that carry them, for instance as a *direct* consequence of antibiotic consumption or *indirectly* due to environmental selection or host factors (e.g., prevalent disease) that impact microbial community composition; 2) *external transmission* via antimicrobial resistance gene carrying bacteria (ARBs). We associated resistome diversity, composition, and the overall ARG load with various socio-demographic parameters - some of which can be used as proxies for the above ecological processes - as well as long-term morbidity and mortality risk.

The World Health Organization has called for research on the demographic parameters underlying the AMR emergence and spread in human populations^14^ to tackle the AMR crisis effectively. Previous studies based on country-level statistics have shown that antibiotic resistance varies with socioeconomic markers^10,15,16^ and antibiotic use^6,17^. However, few studies have analyzed participant-level variation and the role of demographic and lifestyle factors in shaping the resistome within specific populations. Moreover, the associations between population-level resistome variation and long-term mortality and morbidity risk remain largely uncharacterized.

We investigated factors associated with gut resistome variation, as well as prospective mortality and morbidity risk associated with ARG load in a well-characterized Finnish population cohort of 7,095 adults across six regions of Finland with varying demographics (mean age 49, 55% women). The cohort represents the general, non-institutionalized population without acute infections (Figure 2; FINRISK^1^). Based on metagenomic profiles from fecal samples collected in 2002, we studied gut resistome composition and diversity, the total ARG load, and the factors contributing to their observed variation. We defined the total ARG load as the cumulative relative abundance of all observed ARGs. Data on address-level geographic location, diet, household income level, prescription drug purchases, diseases, and causes of death until 2019 were gathered from electronic population registers, health examinations, and complementary questionnaires (see *<u>Methods</u>* for a description of data sources and covariate selection including analysis of collinearity in key covariates). Summary statistics for all antibiotics purchases during the seven years before sampling are provided in <u>Extended Data Table 1</u>. We combined these data to investigate how geographic and demographic factors, lifestyle, and health shape the antibiotic resistome, and what the taxonomic underpinnings of these associations are. Moreover, we investigated the prognostic value of the resistome for mortality due to infectious or other causes. These analyses shed light on the putative ecological and epidemiological patterns of antibiotic resistance at the population level and their health implications.

**Figure 2:**
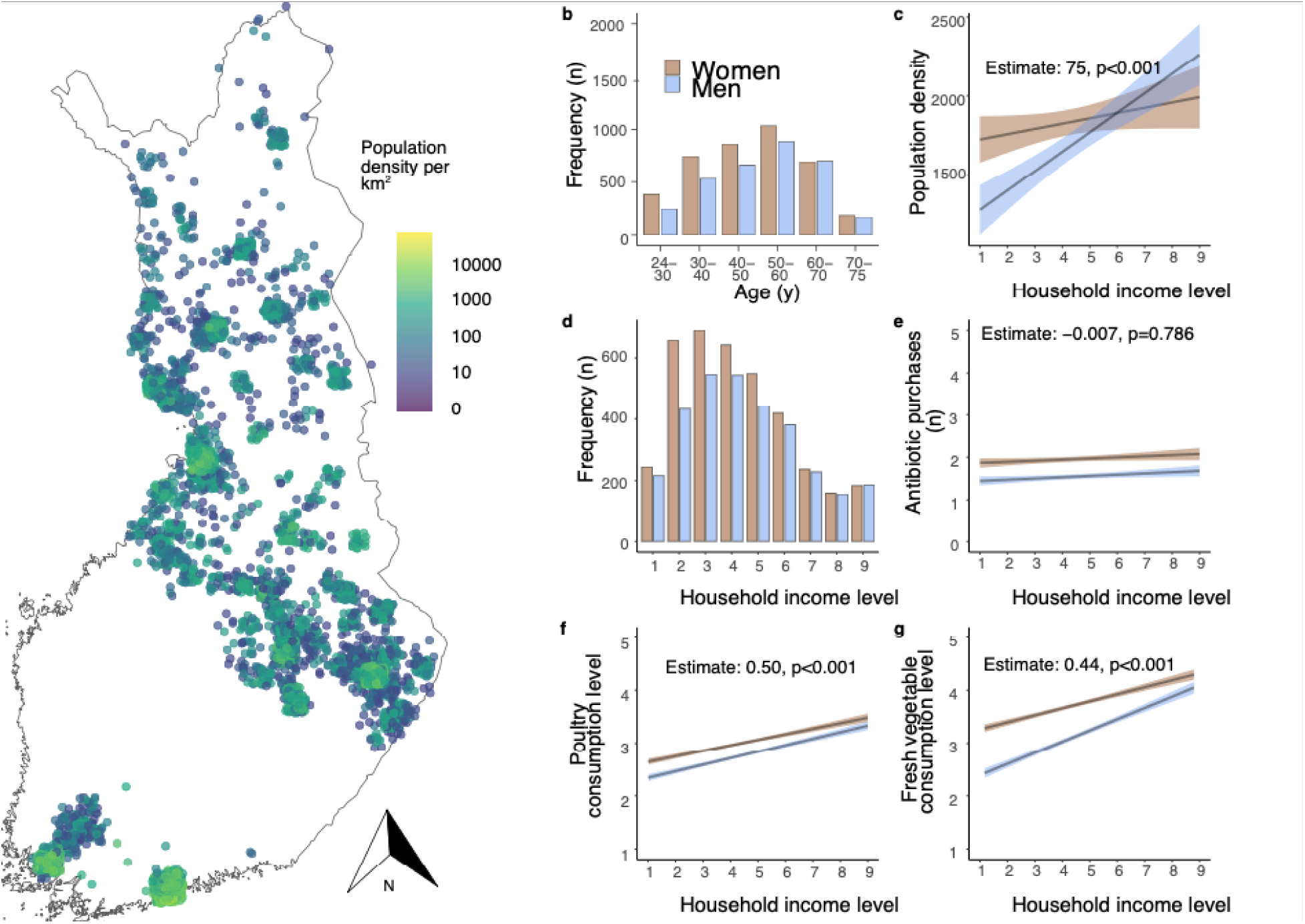
Overview of the FINRISK cohort (N=7,095) **a** Geographical distribution of the cohort participants. The jittered data points indicate the population density per km2. Some individual points at remote locations were removed to obscure participants’ addresses. **b** Age distribution by decade, shown for males (M; blue) and females (F; brown) **c** Population-density versus household income level (scale 1-9). **d** Household income level (scale 1-9) **e** Household income level versus antibiotic purchases during the seven years before the sample collection. **f** Household income level versus poultry consumption (scale 1-5, from less than once a month to multiple times per day), and **g** fresh vegetable and salad consumption. Panels c, e, f, and g include linear models’ estimated slope and p-value. See *<u>Methods</u>* for a more detailed description of the scales.

## RESULTS

### Predictors of ARG load and diversity

The ability to predict ARG load and other resistome features could help identify individuals at risk and derive population-wide estimates of resistome variation (Figure 3). We trained a supervised machine learning model (boosted GLM; *<u>Methods</u>*) to predict ARG load from gut microbiome composition and demographic, health, and lifestyle factors. Through cross-validation on left-out (test) data, we identified key predictors of ARG load and quantified their ability to provide generalizable predictions on new individuals (Figure 3b).

**Figure 3:**
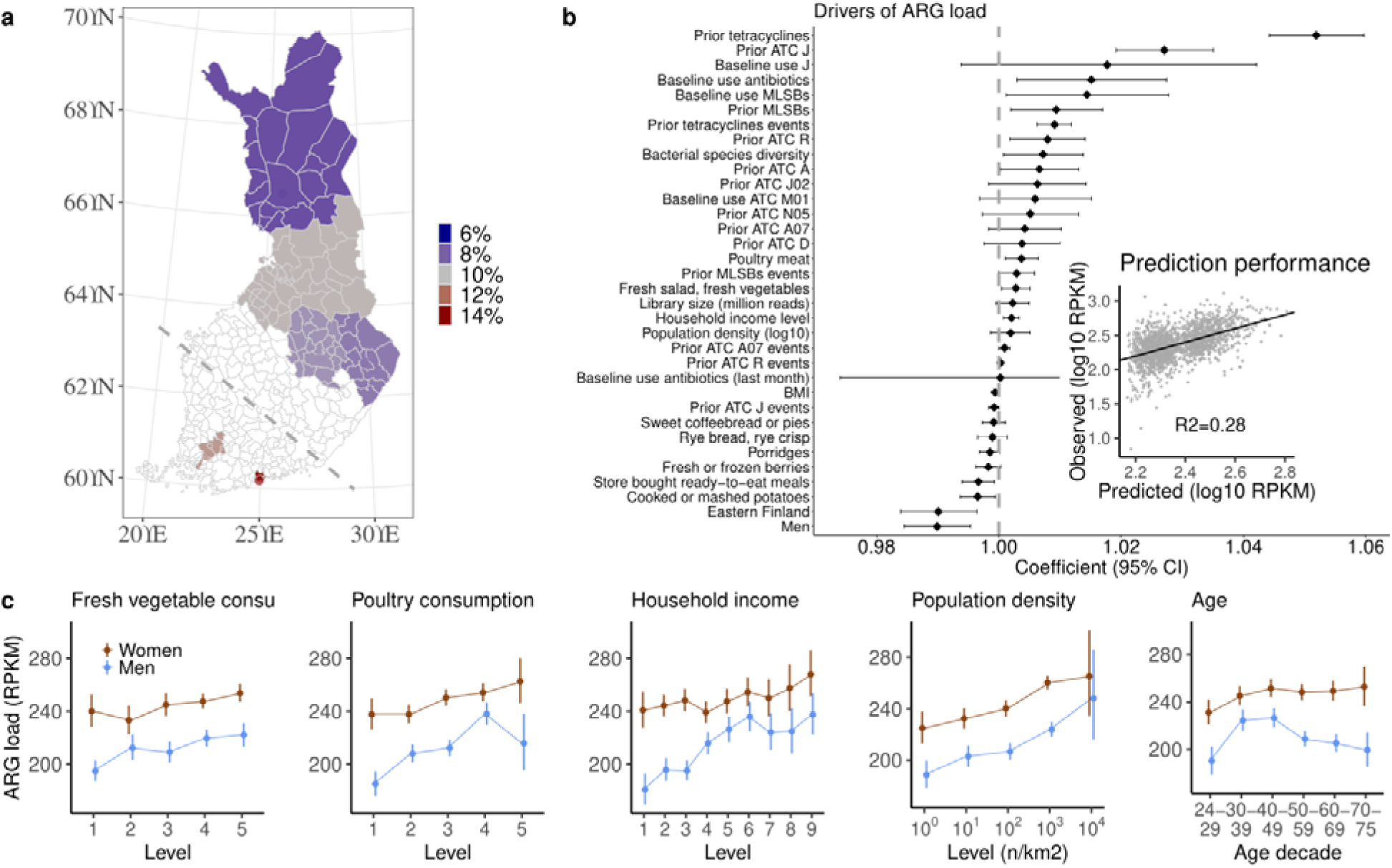
ARG load prediction and variation. **a** Regional variation in total ARG load across the six regions in the FINRISK cohort (North Karelia and North Savonia in the east, Oulu region and Lapland in the north, Turku region in the southwest and Helsinki capital region in the south). The color indicates the fraction of the population in the high ARG group for each region (>458 RPKM; the sum of all normalized ARGs per kilobase per million reads, i.e., the top-10% quantile in this cohort; <u>Extended Data Table 3</u>). The dashed line illustrates the East-West split (the Turku and Helsinki regions in the West and the other four regions in the East). **b** Drivers of ARG load (boosted GLM effect size; see *<u>Methods</u>*). For drivers of resistome diversity, see <u>Figure S1a</u>. The line plot shows each predictor’s effect sizes and 95% confidence interval. The inset shows the predicted and observed ARG load in test data (R2 = 0.28). *Prior X*: purchase of drug X in the seven years before sampling (yes/no), *Prior X events*: number of purchases of drug X in the seven years before sampling, *Baseline X*: Purchase of drug X in the six months before sampling (yes/no). ATC class abbreviations: *A* = Alimentary tract and metabolism, *A07* = Antidiarrheals, intestinal anti-inflammatory/anti-infective agents, *D* = Dermalogicals, *J* = Anti-infectives, *J02* = Antifungals, *M01* = Anti-inflammatory and antirheumatic products, *N05* = Psycholeptics, *R* = Respiratory system. **c** Association between ARG load and fresh vegetables and poultry consumption, household income, population density, and age (N=7,095: see *<u>Methods</u>* for variable descriptions and <u>Supplementary Table 3</u> for the numerical estimates). Similar trends can be observed within individual regions (<u>Figure S2)</u>.

The prediction model could explain 28% and 21% of the variation in ARG load and resistome diversity, respectively, in the independent test set (Figure 3b; <u>Figure S1a</u>). Key predictors for both included antibiotic and drug use, diet, gender, household income, East/West geographic division, population density, and bacterial species diversity (see also Figure 3c). Similar, albeit sometimes non-significant trends could be observed independently within each of the six geographical regions (<u>Figure S2</u>, <u>Extended Data Tables 2-3</u>). Including bacterial families as predictors increased the explained variance to 32% in the test set (boosted GLM, <u>Figure S1b</u>), suggesting that microbial community composition contributes to the ARG load on top of the other covariates. Antibiotic use explained the largest proportion of variance (GLM with just the respective covariates, 27%) in ARG load in the test data, followed by prevalent bacterial families (3%), demographic variables (household income and gender; 2%), geography (East/West; 1%) and diet (1%). Resistome diversity was primarily explained by species diversity (11%) and antibiotic use (6%), followed by demographics (gender and household income) (3%), geography (1%), population density (1%), and diet (1%) (results on ARG diversity are shown in <u>Figure S1a</u>). Antibiotic use, diet, population density, geography, and demographic factors represent proxies for the selection and transmission of antibiotic resistance genes (see Figure 1). Our data confirms they have substantial predictive value on individual ARG load and diversity.

### Drug use

Antibiotic use promotes the selection of antibiotic resistance. The majority of variation in ARG load was explained by antibiotic use alone. Prior use of several classes of antibiotic and non-antibiotic therapeutics listed in the Anatomical Therapeutic Chemical Classification System (ATC) during the past seven years was positively associated with ARG load (p < 0.05, <u>Supplementary Table 1</u>). Consumption of antimicrobials (ATC class J) was associated with a 55% higher ARG load (p < 0.001) compared to no antimicrobial exposure in the past seven years.

In particular, prior tetracycline (ATC class J01) purchases were associated with a 67% increase in ARG load (p < 0.001), and MLSB antibiotics with a 39% increase (ATC class J01F; p < 0.001). These associations remained robust after controlling for other participant-level data, such as diet (Figure 3b). Tetracycline was the second most frequently purchased antibiotic class in the cohort (<u>Extended Data Table 1)</u> and has been widely used since the 1950s. It is also very commonly used in animal production^18^. Moreover, purchases of respiratory medication (ATC class R) were associated with a higher ARG load after controlling for past antibiotic use, demographic factors, and other participant-level data (Figure 3b). Other associations between non-antibiotic drugs and ARG load were not robust to adjusting for other covariates or for abundances of bacterial families (<u>Figure S1</u>). Although some of these drugs might have selective effects favoring resistance, drug use can also be a proxy for additional, unobserved covariates. These observations highlight the need to consider the long-term impact of the use of antibiotics.

### Diet

Food could directly contribute to antibiotic resistance by transmitting ARGs to the gut, for instance, by consuming fresh produce and meat^13,19^. Another way food could influence antibiotic resistance is through indirect selection, that is, by promoting or inhibiting the growth of ARB. For instance, it has been hypothesized that high-fat, high-sugar, and processed foods could promote the growth of Proteobacteria^20^ or other bacterial taxa that tend to carry more ARGs. Conversely, high-fiber foods could select against ARB^21^. Systematic population-level data associating specific foods and antibiotic resistance is limited^21,22^, and the relative importance of selection and transmission in food-mediated resistance is yet to be characterized.

We associated ARG load and diversity with the self-declared habitual consumption frequency of 42 food groups (*<u>Methods</u>*). Poultry had the strongest positive association with ARG load (4% average increase per consumption level, adjusted for antibiotics; p < 0.001; <u>Supplementary Table 1</u>), followed by fresh vegetables and salad (3% increase per level; p < 0.001). In contrast to the other dietary components, the associations remained robust for poultry and fresh vegetables even after controlling for other covariates, including socio-demographics and antibiotic use (p < 0.05 Figure 3b-c). Moreover, their association with ARG load remained significant also when controlling for the abundance of bacterial families (<u>Figure S1</u>, p < 0.05), which suggests that this effect is not due to indirect selection. These results support the hypothesis that these foods contain ARBs that can transfer to the gut.^23,24^ We did not find a similar association for beef or pork, which is in line with the fact that other meat production animals in Finland were reported to have much less antibiotic resistance than poultry at the time of sampling^25^.

In contrast, highly processed, high-sugar, and high-fat foods did not exhibit robust associations with ARG load and diversity. We observed weak associations with ARG load for dietary components such as chocolate and fast food. However, they were not robust to adjusting for participants’ covariates (1% increase per level, p = 0.05-0.15; Figure 3b). Nevertheless, the link between these foods and ARG load and diversity may be partially confounded by antibiotic exposure and diseases, which covary with diet and likely influence the abundance of ARG-containing taxa proposed to respond to high-fat diets, such as Proteobacteria^20^. Interestingly, some of these foods were, in fact, inversely associated with ARG load, as were also total cholesterol and BMI (p < 0.05). This might be partly explained by the lack of fresh vegetables and poultry in the typical diets containing these foods. Associations with high-fiber foods were mixed: both fresh and cooked vegetables were positively associated with ARG load, whereas berries and rye bread had an inverse association (p < 0.05). Taken together, our findings suggest that the influence of population-level dietary variation on antibiotic resistance is dominated by transmission from food rather than indirect selection by diet.

### Population density and urban regions

Geography is a key epidemiological parameter that co-varies with many genetic, demographic, and lifestyle factors that can contribute to the emergence and spread of antibiotic resistance. In Finland, the Eastern and Western populations have well-characterized genetic and lifestyle differences^26^. Whereas earlier large-scale studies have consistently reported geographic variation in antibiotic resistance between countries^5,15,17^, these results have been largely limited to country-level aggregates and thus lack the participant-level resolution that would be necessary for detailed quantification of resistome variation. We used the available participant-level data from national population registers to derive high-resolution estimates of population density based on the participants’ home addresses. This enabled a high-resolution analysis of geographic variation.

Differences in resistome composition between geographic regions were significant in most comparisons (<u>Extended Data Table 4</u>, PERMANOVA, p=0.0015), albeit small (explained variance <0.1%; p<0.002). Eastern Finns had a generally lower ARG load and diversity even after controlling for (family-level) microbiota composition diet, health, population density, and demographic factors (boosted GLM, p > 0.05, Figure 3b; <u>Figure S1</u>). Thus, the variation between East and West could be partly due to other differences, such as limited spatial dispersal, lifestyle covariates, and differences in the genetic background. Urban regions were enriched in individuals with a high ARG load (top-10% quantile; >458 RPKM. This trend was particularly notable around the two urban centers: the Helsinki capital area and Turku, the third-largest urban region in Finland (Figure 3a; <u>Extended Data Table 3</u>). The highest regional ARG load and diversity were observed in these two cities. In contrast, the lowest median ARG load was observed in Lapland, a rural region with a remarkably low population density. Compared to Lapland, the median ARG load was 20% higher in Helsinki and 13% higher in Turku. The increase in the prevalence of high-ARG individuals was even higher (84% and 47%, respectively), suggesting that moderate increases in overall ARG load in urban regions increase the risk of acquiring a high ARG load. This could be explained by increasing ARG transmission in densely populated areas^12^, a hypothesis supported by the observation that ARG load and diversity increased also more generally with population density (Figure 3c; <u>Supplementary Table 1</u>).

### Demographics

Next, we investigated the associations between ARG load and specific socio-demographic factors (Figure 3, <u>Supplementary Table 1</u>). Men had 92% lower average ARG load than women (95% CI 0.89-0.94, p<0.001; log-linear model); the same trend could be observed across all six regions (<u>Figure S2</u>). Women tend to purchase more antibiotics (41% more purchases on average in a linear model adjusted for age; p<0.001) and consume raw vegetables more frequently than men (52%; p<0.001), regardless of household income. Nevertheless, the difference between genders remained significant even after controlling for other covariates, including antibiotics, diet, and the relative abundance of bacterial families (p<0.05, Figure 3b, <u>Figure S1)</u>. Potential causes for this result include differences in occupation or caretaking, prevalence of urinary tract infections^14^, and propensity to seek medical care^27^. Household income was positively associated with ARG load, with a 2% average increase in ARG load per income level (p < 0.001). This association was again robust to differences in antibiotic use and other participant-level covariates, including abundance variations in bacterial families. This finding contrasts with the generally positive association between higher socio-economic status and health^28^, and could be partly explained by lifestyle factors associated with a higher income: these participants consumed more raw vegetables, lived in more densely populated areas (Figure 2, <u>Extended Data Table 1</u>), and are generally more engaged in international travel^29^.

### Resistome composition reflects bacterial phylogeny

Tetracycline resistance was found among virtually all participants (prevalence 100%), whereas the prevalence of other ARG classes was more variable (Figure 4a; <u>Figure S4</u>). Resistome composition and diversity tend to co-vary with bacterial phylogeny^9^ as bacterial taxa differ in their tendency to harbor ARGs^3,30^. In our data, bacterial species diversity partly explained resistome diversity (R2=0.11; Figure 4b; <u>Extended Data Fig 1a</u>), although association with ARG load remained weak (R2=0.001, Figure 3b). We observed phylogenetic clustering among some of the most prevalent ARGs (Figure 4d). The tetracycline resistance genes covered a broad phylogenetic range, which could partly explain their high prevalence. Macrolide resistance gene *ermB* was observed in a more limited yet phylogenetically disjoint set of taxa (e.g., in *Bifidobacterium*, *Clostridium*, and *Faecalibacterium*). *Escherichia* and *Klebsiella* carried the largest number of unique ARGs. Abundant ARGs were often found in commensal taxa: for instance, beta-lactam resistance gene *cfxA6* was detected in *Bacteroides*, *Parabacteroides*, *Veillonella*, and *Prevotella* (BLAST *nr* database^31,32^, Figure 4d). To further examine co-variation between resistome and microbiota composition, we identified five sub-communities, or *enterosignatures* (ES)^33^ of co-varying bacterial genera (<u>Figure S6</u>), which collectively explained 82% of all genus-level variation. They were respectively dominated by members of Bacteroides (ES-*Bact*), Firmicutes (ES-*Firm*), Prevotella (ES-*Prev*), Bifidobacterium (ES-*Bifi*), and Escherichia (ES-*Esch*). Each ES was associated with a different characteristic resistome profile (Figure 4c, <u>Extended Data Table 5)</u>. For instance, ES-*Prev* was associated with high beta-lactam and low tetracycline resistance gene abundances. High ES-*Bact* and ES-*Esch* and low ES-*Prev* and ES-*Bifi* were associated with an increased ARG load. The analysis suggests a non-monotonic relation between ES abundance and ARG enrichment for ES-*Bact*, ES-*Firm,* ES-*Bifi,* and ES-*Esch*, indicating potentially complex ecological relations between bacterial community composition and antibiotic resistance. These observations are supported by the analysis of individual bacterial families (<u>Figure S5</u>; <u>Supplementary Table 1</u>). Overall, these findings emphasize the role of microbiota composition as a key confounder in population-level analyses of resistome variation.

**Figure 4.**
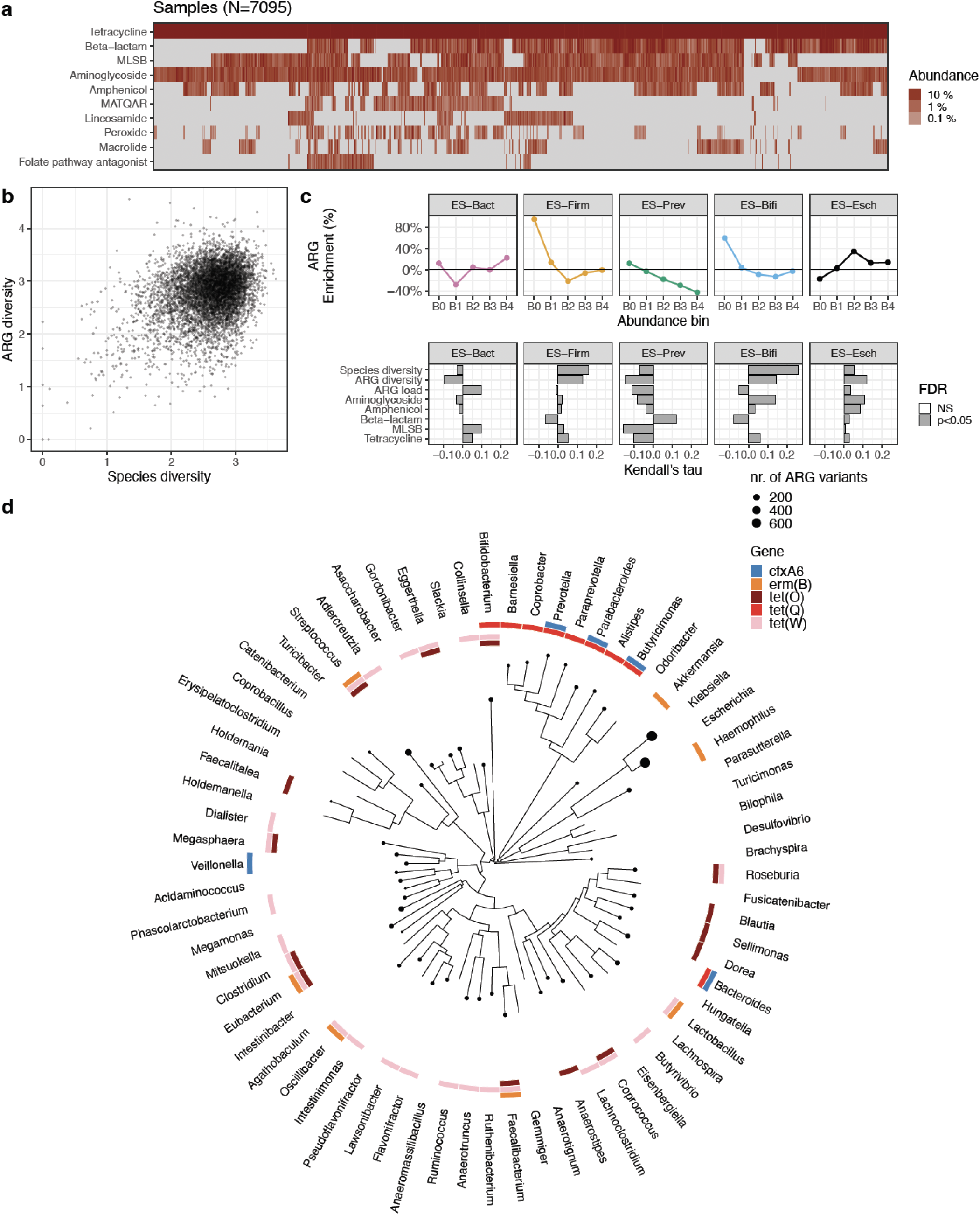
Resistome variation and taxonomic composition. **a** Resistome composition by the relative abundance for the 10 most abundant ARG classes in the FINRISK cohort. **b** Bacterial species diversity and resistome diversity (Shannon index; Pearson r=0.32). **c** *Top panel*: High ARG load (>458 RPKM) enrichment by enterosignature (ES) abundance (B0: not detected; B1-B4: 25% abundance quartiles); each ES represents a sub-community of co-varying genera (<u>Figure S6</u>). *Lower panel* indicates associations between each ES, bacterial species diversity, resistome diversity, total ARG load, and the five most dominant ARG classes (Kendall’s Tau; <u>Supplementary Table 2</u>). The gray bars indicate significant associations (p<0.05). Abbreviations: MLSB: “Macrolide, Lincosamide, Streptogramin B”; MATQAR: "Macrolide, Aminoglycoside, Tetracycline, Quinolone, Amphenicol, Rifamycin". **d** Phylogenetic relatedness among the most prevalent genera in the FINRISK cohort and their association with the most prevalent ARGs. Node size indicates the total number of ARGs found in each genus according to the BLAST *nr* database; the colors indicate a match between each genus and the ARGs.

### Antibiotic resistance is associated with long-term mortality and sepsis risk

We gathered follow-up data on all major health events, including deaths from the baseline sample collection in 2002 until 2019. This data allowed us to link the baseline ARG load to mortality rates and sepsis incidence during the 17 years following sample collection. We anticipated that ARG load could predict all-cause mortality risk because antibiotic-resistant infections may occur as comorbidities and thus contribute to mortality risk due to reduced antibiotic efficacy and as a marker for dysbiotic gut microbiota. In total, 863 (12.2%) of the participants died during the 17-year follow-up period, and 197 (2.8%) got sepsis. We estimated time-to-event associations for selected variables with a probabilistic Cox model (*<u>Methods</u>*). We controlled mortality-associated covariates (age, smoking, gender, diabetes, use of antineoplastic and immunomodulating agents, systolic blood pressure, and self-reported antihypertensive medication) and (log10) relative abundance of *Enterobacteriaceae*. Indeed, this bacterial family is known to harbor many antibiotic-resistant pathogenic members^34^, and we have previously reported an association between its relative abundance and increased risk of all-cause and cause-specific mortality in the same cohort^35^. Furthermore, microbiome composition has been shown to be associated with infection mortality and hospitalizations^36^. In addition, we controlled income and fresh salad and vegetable consumption as potential confounders as they have been inversely associated with mortality^28,37^.

We observed a positive association between ARG load (log10 RPKM) and increased mortality risk (median HR 1.34; Figure 5a). Notably, this association with all-cause mortality was stronger for ARG load than for *Enterobacteriaceae* (median HR 1.08) or systolic blood pressure (median HR 1.04). Moreover, the mortality associated with the high ARG load was more significant in women (Figure 5b). ARG load was specifically associated with increased mortality due to respiratory causes (median HR 2.22; <u>Figure S7)</u>, although the sample sizes for cause-specific events, including infectious mortality other than respiratory infections (ICD-10 A; N=13), remain relatively low (<u>Extended Data Table 7</u>). Moreover, we observed an association between ARG load and an increased sepsis risk (median HR 2.22; <u>Figure S7</u>). The associations between ARG load, all-cause mortality, mortality by respiratory causes, and sepsis remained significant when controlling for the abundance of *Enterobacteriaceae* (<u>Extended Data Tables 6-8</u>), other prevalent bacterial families, or the 42 recorded food groups as covariates (Cox model; p>0.05). These observations suggest that ARG load is a robust risk factor for mortality and sepsis (Figure 5; <u>Figure S7</u>).

**Figure 5:**
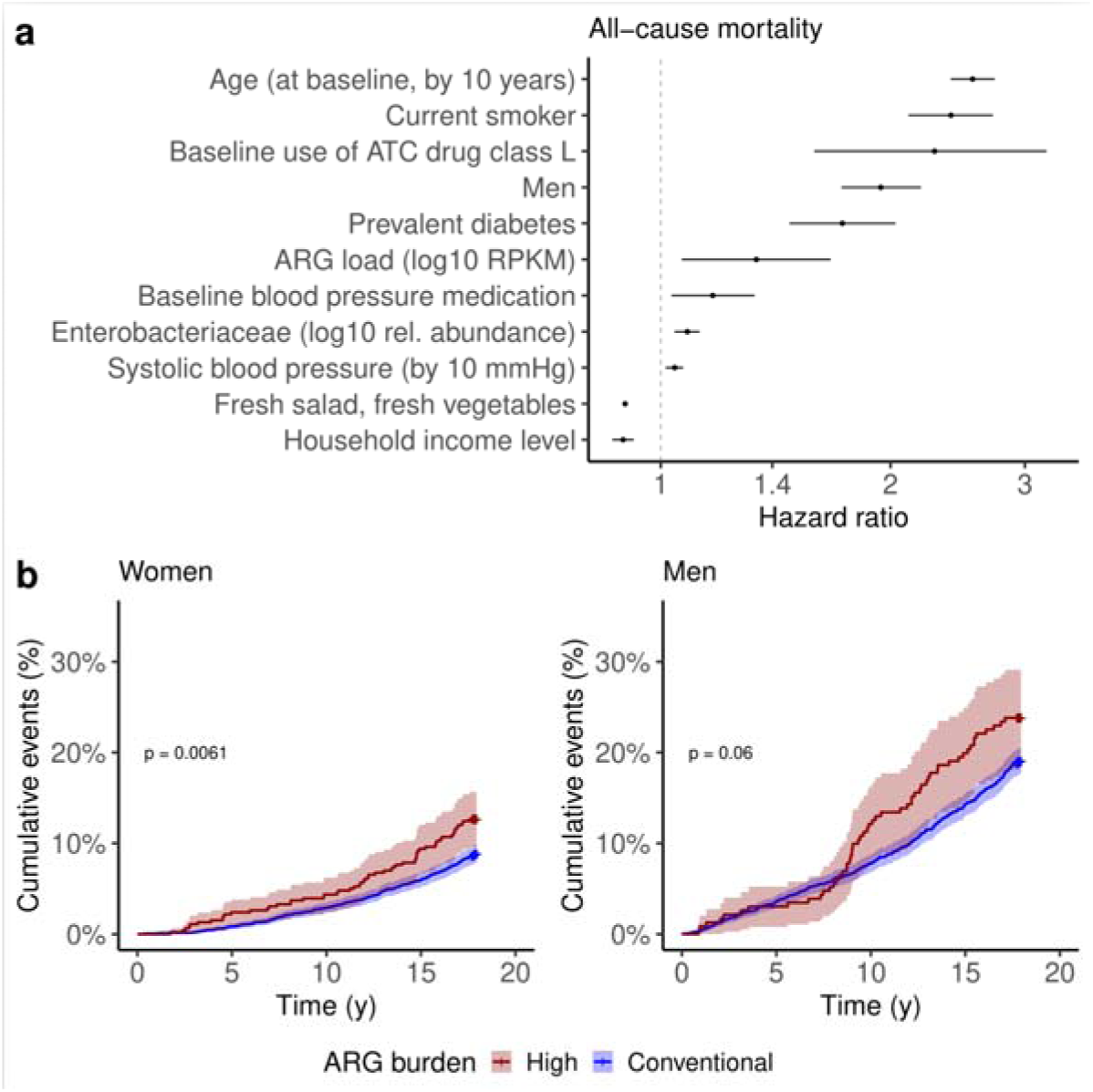
Antibiotic resistance gene (ARG) load predicts long-term mortality risk. **a** Total ARG load associates with long-term mortality risk in a 17-year follow-up (proportional hazards; <u>Extended Data Table 6</u>). The model is adjusted for *Enterobacteriaceae* relative (log10) abundance, age, smoking, gender, diabetes, antineoplastic and immunomodulating agents, body mass index, self-reported antihypertensive medication, systolic blood pressure, recent antibiotics use (six months before baseline), household income, and fresh salad and vegetable consumption. The median hazard ratio (HR) is shown for each variable, along with the 95% credible intervals (CI); variables whose CI overlaps with 1 (no association) are excluded from the graph. **b** Cumulative incidence of all-cause mortality during the follow-up period for individuals stratified by high (red; >458 RPKM) and conventional (blue) ARG load. High ARG load is associated with significantly higher mortality among women (p=0.006; log-rank test; multivariate Cox). A similar but non-significant trend is observed in men (p=0.06). Associations of ARG load with sepsis and cause-specific mortality are shown in <u>Figure S7</u>.

## DISCUSSION

We have characterized the population variation and prognostic potential of gut antibiotic resistome in a single representative cohort of Finnish adults. Our results highlight ecological and epidemiological processes that can facilitate the emergence and spread of antibiotic resistance in human populations. Moreover, our data indicates that ARG load can predict long-term sepsis and mortality risk, with a prognostic potential comparable to blood pressure in a 17-year follow-up. The strongest predictor of ARG load was prior use of antibiotics. This suggests that the *direct selection* of antibiotic resistance genes induced by antibiotic consumption is a key mechanism underlying individual levels of ARG load, potentially several years ahead.

Moreover, further *indirect selection* of the ARGs could be caused by variations in ARG-carrying bacteria in the microbiome, for instance, due to differences in immune function or lifestyle. Finally, we found that known proxies for ARG *transmission* - high population density, income level, and certain food groups - could predict individual ARG load. Thus, these findings highlight population-level patterns that may contribute to antibiotic resistance selection and transmission.

Each study must be evaluated in the context of its limitations. Sequence-based identification of ARGs is not a guarantee of phenotypic antibiotic resistance of bacteria but rather a proxy for antibiotic resistance potential in a microbial community^3^. The patterns observed in a single population may not be generalizable to other countries. Additionally, the lack of longitudinal metagenome data limits our ability to assess causal relations. Our analysis is based on shallow, short-read metagenome profiling. This emphasizes the most abundant and prevalent ARGs and does not allow for the assembly or the creation of draft genomes, which would allow for more detailed information about gene variants as well as the context of the ARGs. The limited sequencing depth is balanced by the sample size, which provides sufficient statistical power for generalizable predictions and distinguishing between the effects of multiple potential confounders.

This is the largest currently available population study quantifying the participant-level variation of antibiotic resistance and its socio-demographic determinants. Our results confirm the need to consider such factors and stratify individuals accordingly when studying resistome variation^14^. Our findings suggest that antibiotic resistance could be a ‘disease of affluence’, as it is more prevalent among high-income individuals with longer life expectancies. Similarly, a previous report found that higher gross domestic product (GDP) per person is linked to higher clinical isolate resistance across countries when controlling for the level of water-sanitation infrastructure, even though not controlling for infrastructure leads to the opposite trend of lower-income countries harboring higher resistance^10,15^. The elevated ARG load among women, high-income participants, and the urban population calls for studies on demographic differences in morbidity and mortality associated with antibiotic resistance.

Interestingly, despite their generally higher ARG load, the overall mortality rates are generally lower in women^27^ and urban high-income groups^28^. These groups also had a lower risk of acquiring sepsis and dying of respiratory causes. The risks associated with a higher ARG load might be thus mitigated by other factors in these subpopulations. However, the observed prognostic potential of high ARG load and the increasing number of reported AMR-related deaths^38,39^ implies that this might be changing. Resistome composition could complement the ongoing efforts to define a healthy microbiome^40^.

Antimicrobial resistance and infection mortality^38^ are rising as leading causes of mortality globally. This trend may be exacerbated by shifts in lifestyle towards conditions that favor ARG selection and transmission through increased consumption of antibiotics and animal protein, urbanization, and international travel. In Finland, poultry consumption has increased by 57% during the past two decades^41^ since our original sample collection, and the population has become more centered in urban areas. Although in Finland, antibiotic use in humans declined over that period, and resistance levels mostly remained stable^42^, global antibiotic consumption and mortality attributed to antibiotic resistance steadily increased over time^43,44^. Moreover, the demand for animal protein and the use of antibiotics in meat production^18,45^ has increased globally. It has been predicted that AMR infections will be the most common cause of death by 2050, marking a shift from cancer and cardiovascular diseases^2^. Our findings represent early warning signals of these shifts.

## STAR Methods

### RESOURCE AVAILABILITY

#### Lead contact

Further information and requests for resources should be directed to and will be fulfilled by the lead contacts: Katariina Pärnänen and Leo Lahti

#### Materials availability

This study did not generate new unique reagents

#### Data and code availability

The metagenomic data are available from the European Genome-Phenome Archive (accession number <u>EGAD00001007035</u>). The phenotype data contain sensitive information from healthcare registers and are available through the THL biobank upon submission of a research plan and signing a data transfer agreement (https://thl.fi/en/web/thl-biobank/for-researchers/application-process).

The analysis source code is available for review as a tar archive. The code will be made public in a GitHub repository with a permanent DOI upon acceptance.

### EXPERIMENTAL MODEL AND STUDY PARTICIPANT DETAILS

The FINRISK population surveys were conducted every five years from 1972 to 2012 with the primary objective of tracking trends in cardiovascular disease risk factors in the Finnish adult population. The FINRISK 2002 study utilized a stratified random sampling approach of individuals between the ages of 25 and 74 from specific regions of Finland (Figure 2). These areas included North Karelia in the east, Northern Savonia in the east, Oulu in the northwest, the province of Lapland in the north, Turku and Loimaa regions in the southwest, and the cities of Helsinki and Vantaa capital region in the south. In addition, we used the West-East split of the regions based on the broad demographic and genetic characteristics of the Finnish population; the Western subset covers the regions of Turku/Loimaa and Helsinki/Vantaa, and the Eastern subset covers the rest of the regions (North Karelia, Northern Savonia, Oulu, Lapland). The sampling procedure was stratified by sex, region, and 10-year age group, resulting in 250 participants in each stratum. For Northern Karelia, Lapland, and the cities of Helsinki and Vantaa, the strata of 65-74-year-old men and women were also sampled, each with 250 participants. The initial population sample comprised 13,500 individuals (excluding 64 who had died or moved away between sample selection and the survey), with an overall participation rate of 65.5% (n = 8,798). Of the participants, n□=□7,231 individuals successfully underwent stool shotgun sequencing. Of those,129 participants withdrew their consent from the THL Biobank at the time of the study. We excluded four individuals with zero reads mapping to the ARG database from the analysis. Subsequently, n□=□7,095 participants (mean age 49 years, 55 % women) remained for unsupervised analysis. Due to a lack of external cohorts with sufficient microbiome profiling and long-term health data, we used two internal subsamples to achieve a 70/30 train test split (n = 5,000 and n = 2,098). We used cross-validation to examine the robustness of the results within the cohort.

#### Population density

The address-level coordinates of the participants were mapped with the *sf* R package^46^ v. 1.0.9. to a 1 km^2^ population grid from 2005, obtained through the *geofi* R package^47^ v. 1.0.7. of the participants’ home addresses ranged from 1 to 19,175 inhabitants/km^2^ (mean 1,753 inhabitants/km^2^). The most densely populated regions are in Southern and South-Western Finland (in the cities of Helsinki and Turku, respectively). We classified the population density into five levels: (<10) 0-9 inhabitants/km^2^; (<100) 10-99; (<1e3) 100-999; (<1e4) 1000-9999; (<2e4) 10,000-20,000. The data points were randomly displaced within a 5 km x 5 km grid to obscure identifiable addresses in the figures. The figures do not show addresses with a population density of less than 10/km^2^.

*Cumulative number of total antibiotic drug purchases* during the past seven years before baseline varied from 0 to 85 (mean 3.3; Figure 2).

*Household income* data was collected based on a questionnaire and was used as the primary demographic descriptor variable alongside gender and age. We also used education level (educational years adjusted for birth year, with the levels low, medium, and high) in the models.

#### Ethical approval

The study protocol of FINRISK 2002 was approved by the Coordinating Ethical Committee of the Helsinki and Uusimaa Hospital District (Ref. 558/E3/20 1 All participants signed informed consent. The study was conducted according to the World Medical Association’s Declaration of Helsinki on ethical principles.

## METHOD DETAILS

### Baseline examination

The FINRISK 2002 survey included a self-administered questionnaire, physical measurements, and blood and stool sample collection. The questionnaire and an invitation to the health examination were mailed to all subjects. Trained nurses conducted physical examinations and blood sampling in local health centers or other survey sites. The participants were advised to fast for ≥4 hours and avoid heavy meals earlier during the day. The venous blood samples were centrifuged at the field survey sites, stored at −70 °C, and transferred daily to the Finnish Institute for Health and Welfare laboratory. Data was collected for physiological measures, biomarkers, and dietary, demographic, and lifestyle factors.

### Stool sample collection

All willing participants were given a stool sampling kit at the baseline examination with detailed instructions. The participants mailed their samples overnight between Monday and Thursday under Finnish winter conditions to the Finnish Institute for Health and Welfare laboratory, where they were stored at −20 °C. The stool samples were transferred frozen in 2017 to the University of California San Diego for microbiome sequencing.

### Stool DNA extraction and library preparation

A miniaturized version of the Kapa HyperPlus Illumina-compatible library prep kit (Kapa Biosystems) was used for library generation. DNA extracts were normalized to 5 ng total input per sample in an Echo 550 acoustic liquid-handling robot (Labcyte I c A Mosquito HV liquid-handling robot (TTP Labtech Inc. was used for 1/10 scale enzymatic fragmentation, end-repair, and adapter-ligation reactions. Sequencing adapters were based on the iTru protocol^48^, in which short universal adapter stubs are ligated first, and then sample-specific barcoded sequences are added in a subsequent PCR step.

Amplified and barcoded libraries were then quantified by the PicoGreen assay and pooled in approximately equimolar ratios before being sequenced on an Illumina HiSeq 4000 instrument to an average read count of ∼900,000 reads per sample.

### Taxonomic and ARG profiling from sequencing data

We analyzed shotgun metagenomic sequences using a pipeline built with the Snakemake^49^ bioinformatics workflow library. We trimmed the sequences for quality and adapter sequences using Atropos^50^. We removed host reads by genome mapping against the human genome assembly GRCh38 with Bowtie2^51^.

We performed taxonomic profiling using MetaPhlAn3^52^ for R1 and R2 reads using the default settings. We mapped the R1 and R2 reads with Bowtie2 v 2.4.4^51^ against the ResFinder database version 2.1.1^53^ with the following options: “-D 20 -R 3 -N 1 -L 20 -i S,1,0 5”. SAMtools v1.10^54^ was used to filter and count reads and if both reads mapped to the same gene the read was counted as one match and if the reads mapped to different genes, both were counted as hits to the respective gene. ARG counts were normalized by library size (number of reads per sample), ARG length, and the sum of all normalized ARGs per kilobase per million reads (RPKM).

### Phylogenetic tree visualization of bacterial taxa and ARGs

We explored the phylogenetic distribution of ARGs in the ResFinder4 database in public sequence data, as our shallow shotgun sequencing did not allow for assigning ARGs to their host genomes using our data. We ran blastn using the ResFinder4 database as the query and the nucleotide collection “nt” database^31^ as a reference, filtering for e-value <10-6 with custom ‘outfmt 6’ including ‘taxid’ for the taxonomic identifier. The blastn results were processed using TaxonKit ^55^ to add genus information based on the identifier to match the genera found in our cohort using genus names. Genera found in MetaPhlAn3 mapping filtered using *mergeFeaturesByPrevalence* function in mia for at least 0.1% abundance in 1% of the samples was used to build a phylogenetic tree of the prevalent genera using ggtree v.3.8.2 ^56^. The most abundant ARGs in the cohort and their presence in the genera were visualized on the tree using the *gheatmap* function from ggtree.

### Register linkage for pre-existing diseases and medication use at baseline

In Finland, each permanent resident is assigned a unique personal identity number at birth or after immigration, which ensures reliable linkage to the electronic health registers. The Finnish health registers cover nearly 100% of all major health events (Hospital Discharge Register, since 1969) and all prescription drug purchases (Drug Purchase Register, since 1995. The quality of the diagnoses in the Finnish national registers has been previously validated^5,6^. Antibiotic drug usage was based on prescription drug purchases (Drug Purchase Register with the Anatomical Therapeutic Chemical (ATC) class J01, which we used as a proxy for actual antibiotic use. Baseline antibiotics use (n□=□1,246) was defined as a purchase with an ATC code of J01* up to 6 months before baseline. The participants were followed through Dec 31, 2019.

Penicillin and other beta-lactam-antibiotics were purchased most often (5390 and 5529 unique purchases in the cohort during 7 years of recording before sampling). Tetracyclines were purchased 5179 times, macrolides, lincosamides, and streptogramins 4620 times. There were no purchases of aminoglycosides (<u>Supplementary Table 1</u>).

### Food questionnaire

Habitual diet was assessed using a food propensity questionnaire (FPQ), which contained 42 food items with choices ranging from 1 – 6 for consumption frequency. Answers denote the following descriptions: An answer 1 ("Less than once a month") 2 ("Once or twice a month") 3 ("Once a week") 4 ("Couple of times a week") 5 ("Almost every day"), and 6 ("Once a day or more often"). For fresh vegetable and salad consumption, the answers 1 and 2 were combined, resulting in new levels 1 (Less than twice a month), 2 (“Once a week”), 3 (“Couple of times a week”), 4 (“Almost every day”), 5 (“Once a day or more often”). For poultry, levels 5 and 6 were combined, resulting in a new level 5 (Almost every day or more often), but the other levels were kept the same as in the original questionnaire. Additionally, the healthy food score ^57^, HFC was used as a proxy for the general healthiness of the diet.

### Regional analysis

For regional analysis, we used the six geographical regions defined above (North Karelia, Northern Savonia, Turku, and Loimaa, Helsinki, and Vantaa, Oulu, and Lapland) or East-West split of the regions.

## QUANTIFICATION AND STATISTICAL ANALYSIS

### Participant data and variable preprocessing

We excluded all variables with near zero variance (*caret* R package^58^ v. 6.0-94) or more than 500 missing values. The ARG load was log_10_ transformed for all the statistical analyses. The dichotomous variables and variables with less than ten levels were unscaled, and other variables, excluding ARG load, were scaled.

### Statistical analysis

All statistical analyses were done in R^59^ version 4.3.1. We corrected for multiple testing using FDR correction (Benjamini–Hochberg) (R *stats* package). We report the adjusted p values. We considered an FDR-corrected P < 0.05 significant. All figures were created with *ggplot2* ^60^ v. 3.4.4 unless otherwise indicated. For all analyses, including microbial taxa, the taxa abundances were centered log-ratio (CLR) transformed to account for compositionality in sequencing data unless otherwise indicated.

### General cohort statistics

ARG load was measured using the total sum of all ARGs’ reads per kilobase per million mapped reads (RPKM). The RPKM values varied considerably among the participants (mean 268 RPKM). The total number of ARG reads mapping to the ARG database had a mean of 468 per sample. There was no association between library size and ARG relative abundances (p = 0.4, linear model). The diversity of ARGs, as measured by the Shannon diversity index, ranged from 0 to 5, with a mean of 3 (Supplementary Figure 1). The number of unique ARGs detected ranged from 1 to 194, with a mean of 42. ARG diversity and load were higher in participants with prior antibiotic use, but ARG richness did not vary significantly (linear model, log-transformed, α = 0.05).

### Alpha diversity

We characterized the alpha diversity of the microbiome with the Shannon index using the complete species-level abundance data for the taxonomic profiles, and using the complete ARG abundance data for the resistome profiles.

### Beta diversity

We used the standard combination of (non-linear) principal coordinate analysis (PCoA) based on the Bray–Curtis dissimilarity index (estimated with the R packages *scater* ^61^ v1.29.4 and *vegan*^62^ v2.6-4 to visualize the overall population variation of the microbiome and resistome composition. The beta-diversity analysis for taxonomic composition was based on species-level relative abundance data from MetaPhlAn3^52^. The beta-diversity analysis for resistome composition was based on the ARG profiles.

### Selection of covariates for modeling

The covariates included in linear models were chosen based on 1148 available covariates. We removed covariates that defined events after sampling and had at least 500 missing values or near zero variance (R package *caret* v6.0-94, *nzv* function with default settings), yielding 134 covariates (<u>Supplementary Table 1</u>) that included prior disease diagnoses for major non-communicable diseases and drug purchase events, as well as geographic region, gender, age, and food frequency. The diseases that passed the filtering criteria included high blood pressure, asthma, diabetes, skeletal fractures, ischemic heart disease, and major cardiovascular events. The majority of diseases did not pass the near-zero variance criterion described above. Two categories for drug purchases were used to investigate if the association differed between recent (6 months) and prior use (7 years).

### Linear models

We performed linear models pairwise with log10(ARG load in RPKM) and ARG diversity and all the explanatory variables that fit the selection criteria. Antibiotic use was controlled using the following parameters: number of events treated with all antibiotics and tetracyclines, prevalent MLSB and tetracycline use, and use of any antibiotic during the past month before baseline. The exponent of the coefficients for ARG load is reported in the main text for ease of interpretation. The pairwise Pearson correlations between key variables (for estimating collinearity) are reported in <u>Extended Data Table 9</u>.

### Boosted GLM

Boosted Generalized linear models (GLMs) with Gaussian distribution were fitted to associate log10(ARG load in RPKM) (and, separately, ARG diversity) with covariates using the R packages *mboost* ^63^ v2.9-8, and *caret* ^58^ v6.0-94. We followed the same selection for boosted GLM as for pairwise linear models, except excluding cholesterol and BMI since they were collinear with income level and diet (<u>Extended Data Table 9</u>), which were of interest (Pearson correlation, p-value <0.05). We further excluded the general diagnosis for mental diseases as it overlapped with drug purchases. Drug purchases were included as both prior (since 1995) and as baseline (past six months) to investigate both short and long-term associations. In boosted GLMs, the generalized linear model is fitted using a boosting algorithm based on component-wise univariate (generalized) linear models. During fitting, the variable selection is performed. The regression coefficients can be interpreted as regular GLM covariates. We reported the exponent of the coefficients for models with log10(ARG load) in the main text for ease of interpretation.

The generalizability of the fitted models was assessed with cross-validation. Five thousand randomly selected participants from the cohort were used for model training, while the remaining 2,095 individuals were used for testing to obtain a 70/30 train-test split. We excluded anthropometric variables such as height and blood markers for cholesterol and triglycerides since those markers are collinear with diet, lifestyle, and gender, which were our main research interests.

Additionally, we fit the boosted GLM by including the most prevalent bacterial families (an abundance of > 0.01% in 1% of samples) and the above-mentioned covariates. Our analysis revealed collinearity between several covariates in the data used to build the boosted GLM (Pearson correlation, p-value <0.05), including income class, sex, and food frequencies for foods such as ready-to-eat meals, raw vegetables, and sausages. Further examination of collinearity is presented in <u>Extended Data Table 9</u>. Despite the collinearity of the covariates, our results show that each model component explains additional variation not captured by the other covariates, and the resulting model does not have variable collinearity.

### GLMs

To validate that the association of fresh vegetables and poultry with ARG load (log10) is not because of underlying taxonomic composition shifts due to the consumption of these foods, we ran pairwise GLMs with fresh salad or poultry consumption and prevalent bacterial families using Gamma distribution and log link. We corrected the p-values with the FDR method.

To validate gender differences, we ran GLMs with fresh vegetable consumption and antibiotic purchases as responses and adjusted with age using Gaussian distribution.

### Probabilistic analyses

To detect potentially non-linear trends in ARG load concerning key covariates (fresh vegetable consumption, poultry consumption, household income, population density, age) and to quantify uncertainties (Figure 3c), we implemented a probabilistic model to predict mean ARG load. We modeled the relation between ARG load and each factor level in the given covariate based on the lognormal distribution using the default values in the R *brm* function from the brms package (version 2.21.0). The model can be summarized in the following pseudocode: brm(ARGload ∼ factor(variable)-1, family = lognormal()). We ran this model separately for each gender. Posterior simulations were used to estimate the mean and credible intervals at the top 10% quantiles for the lognormal distribution at each factor level.

### Survival analysis

We used the Cox-proportional hazards model to predict all-cause mortality during the 17-year follow-up after baseline. We inferred the model parameters with a probabilistic multivariate Cox model using the *brms* v 2.20.4 ^64^ and *tidybayes* v3.0.6 R packages. We used the *ggfortify* ^65,66^ v0.4.16 and *survminer* ^67^ v0.4.9 packages to generate the Kaplan-Meier curve. We verified the probabilistic analyses with frequentist analyses based on the *survival* ^68,69^ v3.5-7 R package. Survival analysis was controlled for *Enterobacteriaceae* abundance, which we previously reported to associate with increasing mortality risk in this cohort (Salosensaari et al. 2021), other mortality-associated covariates used in that publication (age, smoking, gender, diabetes, use of antineoplastic and immunomodulating agents, body-mass index, self-reported antihypertensive medication), and fresh vegetable consumption and income class were included as controls. Income and fresh salad and vegetable consumption were adjusted for in the model since they have been negatively associated with mortality^28,37^ but positively with ARG load. *Enterobacteriaceae* abundance and total ARG load were log10-transformed before the analysis. The median hazard ratio (HR) in Fig. 5a shows a relative change in mortality risk following a unit change in each covariate based on the probabilistic multivariate Cox regression model. The covariates that exhibited strong independent association with mortality are shown (>95% Bayesian credible intervals do not include zero); the credible intervals can be considered a probabilistic version of classical confidence intervals. The Kaplar-Meier curves (Fig. 5b) compare survival between the individuals with high vs. conventional ARG load (the top-10% quantile vs. others; >458 RPKM), controlled for the same covariates as in the Cox regression model. The classical multivariate Cox model further confirms the significant association (p<0.02).

### Enterosignatures

We adapted recently proposed *enterosignatures* to summarize the community composition in a few coherent subcommunities^33^. The enterosignature approach was proposed to complement the earlier attempts to stratify each individual into one of the few distinct community types driven by major groups of gut bacteria. In summary, we applied non-negative matrix factorization (NMF) on the genus-level relative abundances after combining the rare genera (<1% prevalence above 0.1% relative abundance) into a single group (“Other”). We run NMF with 2-10 components with 10 runs using the default parameters in the function *nmf* from the R package NMF (v. 0.26). The Silhouette consensus measure in the function output indicated an optimal solution of 5 NMF components. Frioux et al. (2023) originally reported the same optimal number of components. We observed a notable correspondence of these signatures between FINRISK (<u>Figure S6</u>) and Frioux et al. (2023; Figure 1). Three of the components had the same most abundant genus in both cases (*Bacteroides*, *Prevotella*, *Bifidobacterium*). In the *Firmicutes* component, the most abundant ten genera were *Firmicutes,* which accounted for 94% of this component in FINRISK. The *Escherichia* component was dominated by *Butyrivibrio* in FINRISK, with fewer Proteobacteria in the FINRISK data. Yet, it was the only signature observed in FINRISK associated with *Escherichia* (a scaled component loading 100% for this signature). Notably, despite the differences, including independent data sets, metagenomic preprocessing pipelines, and implementation details, the five ES identified in FINRISK had a direct qualitative correspondence with the initially reported enterosignatures. We also checked that the enterosignature abundances were robust to variations in library sizes (Kendall’s tau; p>0.05 for all ES).

### Kendall’s Rank Correlation

Associations between diversities, ARG load, and bacterial families or Enterosignatures were calculated using Kendall’s Rank Correlation (also known as Kendall’s *Tau*), followed by FDR correction using the *mia* ^70^ package v. 1.9.19 function *getExperimentCrossAssociation*.

## ACKNOWLEDGEMENTS

KP, LL, VL, and GSK were supported by grants from the Research Council of Finland (decisions 348439 to KP; 330887 to LL and VL; 340314 to GSK) and the Alhopuro Foundation (decisions 20220114 to KP and 20210172 to GSK). CGV and GM are supported by an Australian National Health and Medical Research Council (NHMRC) grant GNT2013468. CSC IT Centre for Science computational resources were used for bioinformatic analysis of the samples. BioRender was used to create Figure 1. Shivang Bhanushali is acknowledged for participating in proofreading the manuscript.

## AUTHOR CONTRIBUTIONS

KP wrote the first draft of the manuscript. LL supervised the work. KP, MR, VL and LL analyzed the data. RK, VS, ASH, and TN acquired the data. KP, VS, TN, ASH and LL designed the study. KP, GSK, and LL wrote the manuscript. All authors contributed to and accepted the manuscript.

## DECLARATION OF INTERESTS

R.K. is a scientific advisory board member and consultant for BiomeSense, Inc., has equity, and receives income. He is a scientific advisory board member and has equity in GenCirq. He is a consultant and scientific advisory board member for DayTwo and receives income. He has equity in and acts as a consultant for Cybele. He is a co-founder of Biota, Inc., and has equity. He is a cofounder of Micronoma and has equity and is a scientific advisory board member. The terms of these arrangements have been reviewed and approved by the University of California, San Diego, in accordance with its conflict of interest policies. Other authors declare no competing interests.

