## Supplemental Information for "Population variation and prognostic potential of gut antibiotic resistome"

**Extended Data Table 1: Number of purchases for different antibiotic classes.** Summary of the antibiotic reimbursed purchase events (participants n = 7,095). Data on drug purchases covers seven years before sampling using drug registry data. Antibiotic use is reported using ATC codes for antibiotics. *Purchases* describe the number of reimbursed purchases in the whole data set. *Participants* is the number of participants with reimbursed purchases for the drug. *Mean* and *median* (with range) are the average numbers of reimbursed purchases per person.

|  | **Purchases (nr of events )** | **Participants (N)** | **Mean** | **Standard deviation** | **Median (min-max)** |
| --- | --- | --- | --- | --- | --- |
| Prior antibiotics | 23516 | 5488 | 3.31 | 4.47 | 2 (0-85) |
| Prior non-penicillin betalactams | 5529 | 2667 | 0.78 | 1.34 | 0 (0-19) |
| Prior tetracyclines | 5179 | 2838 | 0.73 | 1.37 | 0 (0-20) |
| Prior penicillins | 5390 | 2971 | 0.76 | 1.44 | 0 (0-38) |
| Prior sulfonamides and trimethoprim | 1581 | 863 | 0.22 | 0.97 | 0 (0-27) |
| Prior MLSBs | 4620 | 2430 | 0.65 | 1.34 | 0 (0-20) |
| Prior quinolones | 1063 | 637 | 0.15 | 0.73 | 0 (0-26) |
| Prior other antibacterials (J01X) | 154 | 87 | 0.020 | 0.27 | 0 (0-9) |

**Extended Data Table 2: Regional variation in ARG load.** For each of the six geographical regions, we show the sample size (N), median ARG diversity, and median ARG load together with the 5% and 95% quantiles, prevalence of the high ARG load individuals (>458 RPKM; calculated based on the top-10% quantile across all regions combined), ratio between the median ARG load between the given region and Lapland, and ratio between the high ARG prevalence between the given region and Lapland. The other regions are compared to Lapland, as this region has the largest sample size, the lowest population density, and the lowest average ARG load.

| **Region** | **N** | **ARG diversity**  median (quantiles) | **ARG load (RPKM)**  median (quantiles) | **High ARG Prevalence** (%) | **Median ARG Ratio** | **High ARG Ratio** |
| --- | --- | --- | --- | --- | --- | --- |
| Lapland | 1462 | 2.8 (6.58-29.85) | 218 (86-529) | 7.5 | 1 | 1 |
| Karelia | 939 | 2.81 (6.63-29.67) | 228 (93-553) | 8.4 | 1.04 | 1.12 |
| Savonia | 1087 | 2.85 (5.91-30.26) | 231 (101-549) | 8.9 | 1.06 | 1.19 |
| Oulu | 1420 | 2.84 (6.99-29.9) | 228 (87-565) | 10.4 | 1.04 | 1.39 |
| Turku | 934 | 2.89 (7.63-31.71) | 246 (111-593) | 11 | 1.13 | 1.47 |
| Helsinki | 1253 | 2.94 (8.57-34.92) | 263 (110-631) | 13.8 | 1.2 | 1.84 |

**Extended Data Table 3: Regional ARG load variation and demographic factors.** Associations between ARG load (log10 RPKM) and the indicated background factors. Linear models fitted separately for each gender (2) and geographical sub-region (6). In addition, results are shown for Eastern and Western Finland; Western Finland covers the urban regions of Helsinki and Turku; Eastern Finland covers the other four regions. The effect size (exponent of the slope fitted on logarithmic ARG load) indicates the relative increase in ARG load per each level of the corresponding variable; the effect size >1 indicates a positive association. Antibiotic use and other covariates have been omitted in these gender- and region-specific models due to low sample sizes and limited power. The analyses for the entire cohort are adjusted for antibiotic consumption and other covariates ([Supplementary Table 1](https://docs.google.com/spreadsheets/u/0/d/1Vn-QHMeFmsuMh8yjpypN9UfioNf-xtgFhaJ-mN7Y2wk/edit)).

| **Variable** | **Region** | **Gender** | **Effect size** | **FDR** |
| --- | --- | --- | --- | --- |
| Baseline age | Lapland | Women | 1.001 | 0.584 |
| Household income level | Lapland | Women | 1.033 | **0.006** |
| Fresh salad, fresh vegetables | Lapland | Women | 1.023 | 0.24 |
| Poultry meat | Lapland | Women | 1.029 | 0.24 |
| Population density (log10) | Lapland | Women | 1.019 | 0.492 |
| Baseline age | Lapland | Men | 0.995 | **0.018** |
| Household income level | Lapland | Men | 1.042 | **0.003** |
| Fresh salad, fresh vegetables | Lapland | Men | 1.04 | **0.05** |
| Poultry meat | Lapland | Men | 1.045 | 0.066 |
| Population density (log10) | Lapland | Men | 1.055 | **0.05** |
| Baseline age | Oulu | Women | 1 | 0.926 |
| Household income level | Oulu | Women | 1.005 | 0.812 |
| Fresh salad, fresh vegetables | Oulu | Women | 1.027 | 0.434 |
| Poultry meat | Oulu | Women | 1.018 | 0.632 |
| Population density (log10) | Oulu | Women | 1.03 | 0.434 |
| Baseline age | Oulu | Men | 0.998 | 0.305 |
| Household income level | Oulu | Men | 1.031 | **0.037** |
| Fresh salad, fresh vegetables | Oulu | Men | 1.032 | 0.138 |
| Poultry meat | Oulu | Men | 1.053 | 0.064 |
| Population density (log10) | Oulu | Men | 1.025 | 0.305 |
| Baseline age | Helsinki | Women | 1.002 | 0.33 |
| Household income level | Helsinki | Women | 0.998 | 0.798 |
| Fresh salad, fresh vegetables | Helsinki | Women | 0.979 | 0.432 |
| Poultry meat | Helsinki | Women | 1.014 | 0.627 |
| Population density (log10) | Helsinki | Women | 1.155 | 0.064 |
| Baseline age | Helsinki | Men | 1.002 | 0.247 |
| Household income level | Helsinki | Men | 1.021 | 0.065 |
| Fresh salad, fresh vegetables | Helsinki | Men | 0.994 | 0.725 |
| Poultry meat | Helsinki | Men | 1.059 | 0.054 |
| Population density (log10) | Helsinki | Men | 1.077 | 0.247 |
| Baseline age | Karelia | Women | 1.003 | 0.42 |
| Household income level | Karelia | Women | 1.004 | 0.88 |
| Fresh salad, fresh vegetables | Karelia | Women | 1.003 | 0.88 |
| Poultry meat | Karelia | Women | 1.033 | 0.42 |
| Population density (log10) | Karelia | Women | 1.025 | 0.65 |
| Baseline age | Karelia | Men | 0.999 | 0.793 |
| Household income level | Karelia | Men | 1.036 | **0.047** |
| Fresh salad, fresh vegetables | Karelia | Men | 1.025 | 0.372 |
| Poultry meat | Karelia | Men | 1.037 | 0.372 |
| Population density (log10) | Karelia | Men | 0.991 | 0.793 |
| Baseline age | Kuopio | Women | 1.003 | 0.222 |
| Household income level | Kuopio | Women | 0.999 | 0.893 |
| Fresh salad, fresh vegetables | Kuopio | Women | 1.028 | 0.214 |
| Poultry meat | Kuopio | Women | 1.034 | 0.214 |
| Population density (log10) | Kuopio | Women | 1.049 | 0.164 |
| Baseline age | Kuopio | Men | 1.002 | 0.47 |
| Household income level | Kuopio | Men | 1.02 | 0.36 |
| Fresh salad, fresh vegetables | Kuopio | Men | 0.982 | 0.47 |
| Poultry meat | Kuopio | Men | 1.038 | 0.36 |
| Population density (log10) | Kuopio | Men | 1.01 | 0.697 |
| Baseline age | Turku | Women | 1 | 0.921 |
| Household income level | Turku | Women | 0.999 | 0.921 |
| Fresh salad, fresh vegetables | Turku | Women | 0.988 | 0.904 |
| Poultry meat | Turku | Women | 0.979 | 0.836 |
| Population density (log10) | Turku | Women | 1.031 | 0.836 |
| Baseline age | Turku | Men | 1.003 | 0.344 |
| Household income level | Turku | Men | 1.014 | 0.344 |
| Fresh salad, fresh vegetables | Turku | Men | 1.021 | 0.344 |
| Poultry meat | Turku | Men | 1.062 | 0.139 |
| Population density (log10) | Turku | Men | 1.031 | 0.344 |
| Baseline age | East | Women | 1.001 | 0.287 |
| Household income level | East | Women | 1.012 | **0.037** |
| Fresh salad, fresh vegetables | East | Women | 1.024 | **0.012** |
| Poultry meat | East | Women | 1.029 | **0.012** |
| Population density (log10) | East | Women | 1.032 | **0.012** |
| Baseline age | East | Men | 0.998 | **0.026** |
| Household income level | East | Men | 1.033 | **<0.001** |
| Fresh salad, fresh vegetables | East | Men | 1.024 | **0.019** |
| Poultry meat | East | Men | 1.046 | **0.001** |
| Population density (log10) | East | Men | 1.027 | **0.042** |
| Baseline age | West | Women | 1.002 | 0.201 |
| Household income level | West | Women | 0.999 | 0.888 |
| Fresh salad, fresh vegetables | West | Women | 0.984 | 0.388 |
| Poultry meat | West | Women | 1.002 | 0.888 |
| Population density (log10) | West | Women | 1.071 | **0.03** |
| Baseline age | West | Men | 1.003 | **0.069** |
| Household income level | West | Men | 1.019 | **0.027** |
| Fresh salad, fresh vegetables | West | Men | 1.005 | 0.698 |
| Poultry meat | West | Men | 1.063 | **0.002** |
| Population density (log10) | West | Men | 1.05 | 0.069 |

**Extended Data Table 4: Dissimilarity in resistome composition between regions.** Dissimilarity between geographical study regions in Finland (PERMANOVA; Bray-Curtis index). Significant FDR-adjusted p-values are bolded.

| **pairs** | **F** | **R2** | **FDR** |
| --- | --- | --- | --- |
| Lapland vs Oulu | 3.0 | 0.001 | **0.005** |
| Lapland vs Helsinki | 26.3 | 0.010 | **0.002** |
| Lapland vs Karelia | 2.5 | 0.001 | **0.010** |
| Lapland vs Kuopio | 4.6 | 0.002 | **0.002** |
| Lapland vs Turku | 14.0 | 0.006 | **0.002** |
| Oulu vs Helsinki | 14.4 | 0.005 | **0.002** |
| Oulu vs Karelia | 2.2 | 0.001 | **0.022** |
| Oulu vs Kuopio | 1.4 | 0.001 | 0.121 |
| Oulu vs Turku | 6.3 | 0.003 | **0.002** |
| Helsinki vs Karelia | 14.7 | 0.007 | **0.002** |
| Helsinki vs Kuopio | 11.5 | 0.005 | **0.002** |
| Helsinki vs Turku | 3.2 | 0.001 | **0.002** |
| Karelia vs Kuopio | 1.5 | 0.001 | 0.121 |
| Karelia vs Turku | 7.7 | 0.004 | **0.002** |
| Kuopio vs Turku | 5.0 | 0.002 | **0.002** |
| Eastern vs Western | 29.5 | 0.004 | 0.001 |

**Extended Data Table 5: Associations between enterosignatures and ARG classes.**

Associations (Figure 4b) between the total (summed) abundance antimicrobial gene classes as detected by *Resfinder* and each enterosignature (Kendall’s rank correlation (tau); FDR correction; no covariates). The ARG load, ARG diversity, and species diversity are also shown. ES-Bact is characterized by *Bacteroides*, ES-Firm by *Firmicutes*, ES-Prev by *Prevotella*, ES-Bifi by *Bifidobacteria*, ES- Esch by *Escheria*. Significant FDR-adjusted p-values are bolded.

| **Variable** | **Enterosignature** | **Tau** | **FDR** |
| --- | --- | --- | --- |
| Amphenicol | ES-Bact | -0.02 | **0.036** |
| Beta-lactam | ES-Bact | 0 | 0.844 |
| MATQAR | ES-Bact | -0.02 | **0.046** |
| MLSB | ES-Bact | 0.1 | **<0.001** |
| Tetracycline | ES-Bact | 0.05 | **<0.001** |
| Amphenicol | ES-Firm | 0.02 | **0.037** |
| Beta-lactam | ES-Firm | -0.06 | **<0.001** |
| MATQAR | ES-Firm | -0.13 | **<0.001** |
| MLSB | ES-Firm | 0.03 | **0.000** |
| Tetracycline | ES-Firm | 0.05 | **<0.001** |
| Amphenicol | ES-Prev | -0.04 | **0.000** |
| Beta-lactam | ES-Prev | 0.12 | **<0.001** |
| MATQAR | ES-Prev | -0.01 | 0.175 |
| MLSB | ES-Prev | -0.16 | **<0.001** |
| Tetracycline | ES-Prev | -0.1 | **<0.001** |
| Amphenicol | ES-Bifi | 0.04 | **0.000** |
| Beta-lactam | ES-Bifi | -0.08 | **<0.001** |
| MATQAR | ES-Bifi | 0.07 | **<0.001** |
| MLSB | ES-Bifi | 0 | 0.919 |
| Tetracycline | ES-Bifi | 0.06 | **<0.001** |
| Amphenicol | ES-Esch | 0.09 | **<0.001** |
| Beta-lactam | ES-Esch | 0.03 | **0.003** |
| MATQAR | ES-Esch | 0.32 | **<0.001** |
| MLSB | ES-Esch | 0.01 | 0.382 |
| Tetracycline | ES-Esch | 0.03 | **0.001** |
| ARG load | ES-Bact | 0.1 | **<0.001** |
| ARG diversity | ES-Bact | -0.1 | **<0.001** |
| Species diversity | ES-Bact | -0.03 | **<0.001** |
| ARG load | ES-Firm | 0 | 0.919 |
| ARG diversity | ES-Firm | 0.14 | **<0.001** |
| Species diversity | ES-Firm | 0.16 | **<0.001** |
| ARG load | ES-Prev | -0.11 | **<0.001** |
| ARG diversity | ES-Prev | -0.14 | **<0.001** |
| Species diversity | ES-Prev | -0.07 | **<0.001** |
| ARG load | ES-Bifi | -0.05 | **<0.001** |
| ARG diversity | ES-Bifi | 0.14 | **<0.001** |
| Species diversity | ES-Bifi | 0.26 | **<0.001** |
| ARG load | ES-Esch | 0.02 | **0.007** |
| ARG diversity | ES-Esch | 0.11 | **<0.001** |
| Species diversity | ES-Esch | 0.06 | **<0.001** |

**Extended Data Table 6: Proportional hazards for total mortality in a 17-year follow-up.** Median effect size and the 95% credible intervals for the probabilistic multivariate Cox proportional hazards model ([Methods](https://docs.google.com/document/d/1z36lt04Dct0uWIWn3eFdyGFqVy88wXPdm9fvUnW7hPQ/edit)).

| **Variable** | **Median HR** | **Quantile 2.5%** | **Quantile 97.5%** |
| --- | --- | --- | --- |
| Age (at baseline, by 10 years) | 2.56 | 2.40 | 2.74 |
| Current smoker | 2.41 | 2.11 | 2.72 |
| Baseline use, ATC drug class L | 2.27 | 1.56 | 3.22 |
| Men | 1.94 | 1.72 | 2.19 |
| Prevalent diabetes | 1.73 | 1.47 | 2.02 |
| ARG load (log10 RPKM) | 1.34 | 1.07 | 1.67 |
| Baseline blood pressure medication | 1.17 | 1.03 | 1.32 |
| Enterobacteriaceae  (log10 rel. abundance) | 1.08 | 1.04 | 1.13 |
| Systolic blood pressure  (by 10 mmHg) | 1.04 | 1.01 | 1.07 |
| Fresh salad, fresh vegetables | 0.90 | 0.86 | 0.94 |
| Household income level | 0.89 | 0.86 | 0.92 |

**Extended Data Table 7: Proportional hazards for cause-specific mortality in a 17-year follow-up.** Median effect size and the 95% credible intervals for the probabilistic multivariate Cox proportional hazards model ([Methods](https://docs.google.com/document/d/1z36lt04Dct0uWIWn3eFdyGFqVy88wXPdm9fvUnW7hPQ/edit)).

| **Cause** | **Median HR** | **Quantile 2.5%** | **Quantile 97.5%** | **Events (N)** |
| --- | --- | --- | --- | --- |
| Gastrointestinal | 2.72 | 0.95 | 8.28 | 42 |
| Respiratory | 2.52 | 1.29 | 5.08 | 103 |
| Cancer | 1.46 | 0.92 | 2.34 | 221 |
| Cardiovascular | 1.45 | 0.95 | 2.28 | 251 |
| Neurological | 1.41 | 0.59 | 3.64 | 58 |
| All | 1.34 | 1.07 | 1.68 | 947 |
| Physical trauma | 1.13 | 0.49 | 2.69 | 64 |

**Extended Data Table 8: Proportional hazards for incident sepsis in a 17-year follow-up.** Median effect size and the 95% credible intervals for the probabilistic multivariate Cox proportional hazards model ([Methods](https://docs.google.com/document/d/1z36lt04Dct0uWIWn3eFdyGFqVy88wXPdm9fvUnW7hPQ/edit)).

| **Variable** | **Median HR** | **Quantile 2.5%** | **Quantile 97.5%** |
| --- | --- | --- | --- |
| Baseline use of ATC drug class L | 2.90 | 1.33 | 5.54 |
| ARG load (log10 RPKM) | 2.22 | 1.33 | 3.65 |
| Age (at baseline, by 10 years) | 1.99 | 1.73 | 2.29 |
| Prevalent diabetes | 1.99 | 1.45 | 2.70 |
| Baseline blood pressure medication | 1.64 | 1.27 | 2.09 |
| Current smoker | 1.63 | 1.21 | 2.19 |
| Men | 1.50 | 1.16 | 1.92 |
| BMI | 1.04 | 1.01 | 1.06 |
| Household income level | 0.91 | 0.85 | 0.97 |

**Extended Data Table 9: Variable correlations**. Reported r and p-values in parentheses for the pairwise Pearson correlation of the covariates used in Fig. 2; calculated using the *rcorr* function from the *Hmisc* R package. See Methods for more detailed descriptions of the covariates.

|  | **Population density (log10)** | **Fresh salad, fresh vegetables** | **Household income level** | **Baseline age** | **Prior antibiotics events** | **BMI** | **Cholesterol** |
| --- | --- | --- | --- | --- | --- | --- | --- |
| Population density (log10) |  | 0.12 (0.000) | 0.06 (0.000) | -0.04 (0.002) | 0.02 (0.115) | -0.09 (0.000) | -0.07 (0.000) |
| Fresh salad, fresh vegetables | 0.12 (0.000) |  | 0.24 (0.000) | -0.01 (0.607) | 0.03 (0.007) | -0.08 (0.000) | -0.06 (0.000) |
| Household income level | 0.06 (0.000) | 0.24 (0.000) |  | -0.15 (0.000) | -0.01 (0.390) | -0.10 (0.000) | -0.02 (0.045) |
| Baseline age | -0.04 (0.002) | -0.01 (0.607) | -0.15 (0.000) |  | 0.03 (0.006) | 0.26 (0.000) | 0.25 (0.000) |
| Prior antibiotics events | 0.02 (0.115) | 0.03 (0.007) | -0.01 (0.390) | 0.03 (0.006) |  | 0.08 (0.000) | -0.06 (0.000) |
| BMI | -0.09 (0.000) | -0.08 (0.000) | -0.10 (0.000) | 0.26 (0.000) | 0.08 (0.000) |  | 0.15 (0.000) |
| Cholesterol | -0.07 (0.000) | -0.06 (0.000) | -0.02 (0.045) | 0.25 (0.000) | -0.06 (0.000) | 0.15 (0.000) |  |

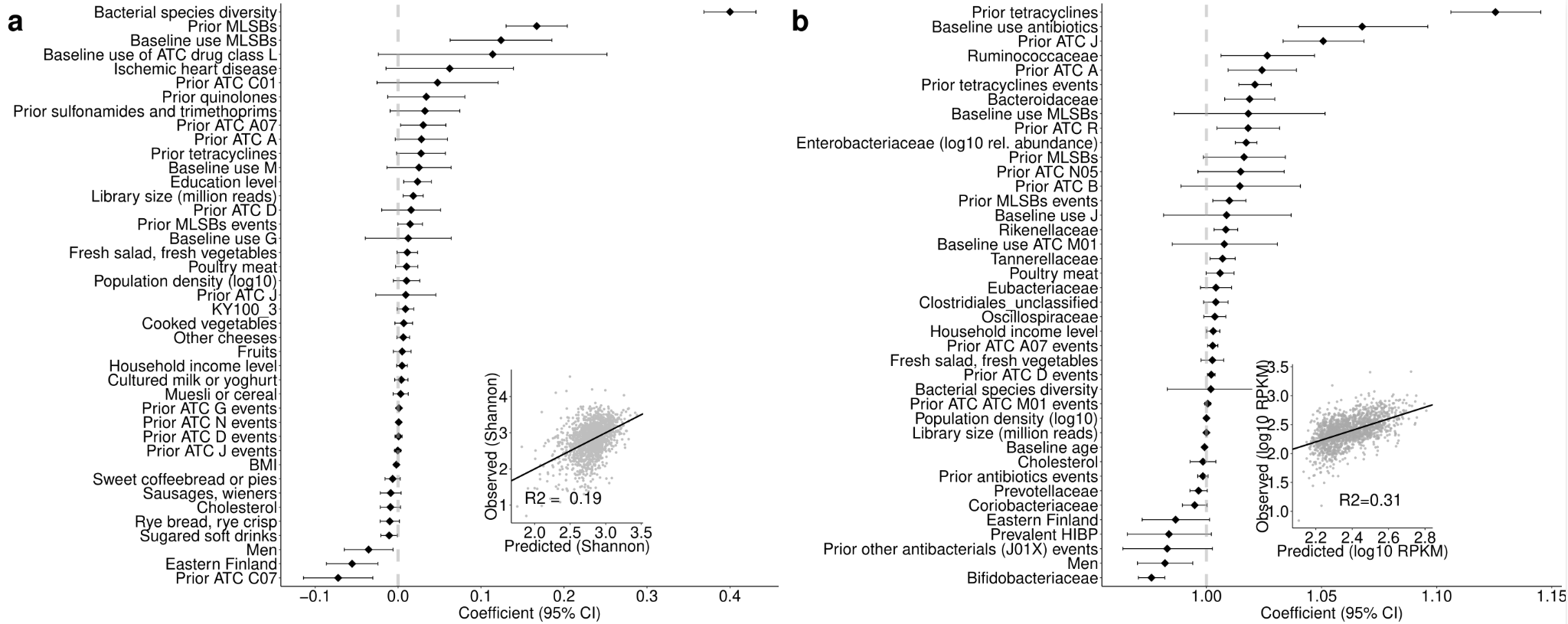

**Figure S1: Drivers of ARG diversity and ARG load a** Drivers of ARG diversity (boosted GLM for ARG Shannon diversity). The line plot shows the estimated effect sizes and 95% confidence intervals for the predictor variables. Inset: Predicted and observed ARG diversity in the leave-out test data. **b** Drivers of ARG load, including bacterial families, exponent of coefficient shown (boosted GLM for log10 ARG load). Bacterial abundances are indicated as log10 relative abundance. Inset: Predicted and observed ARG load in the leave-out test data.

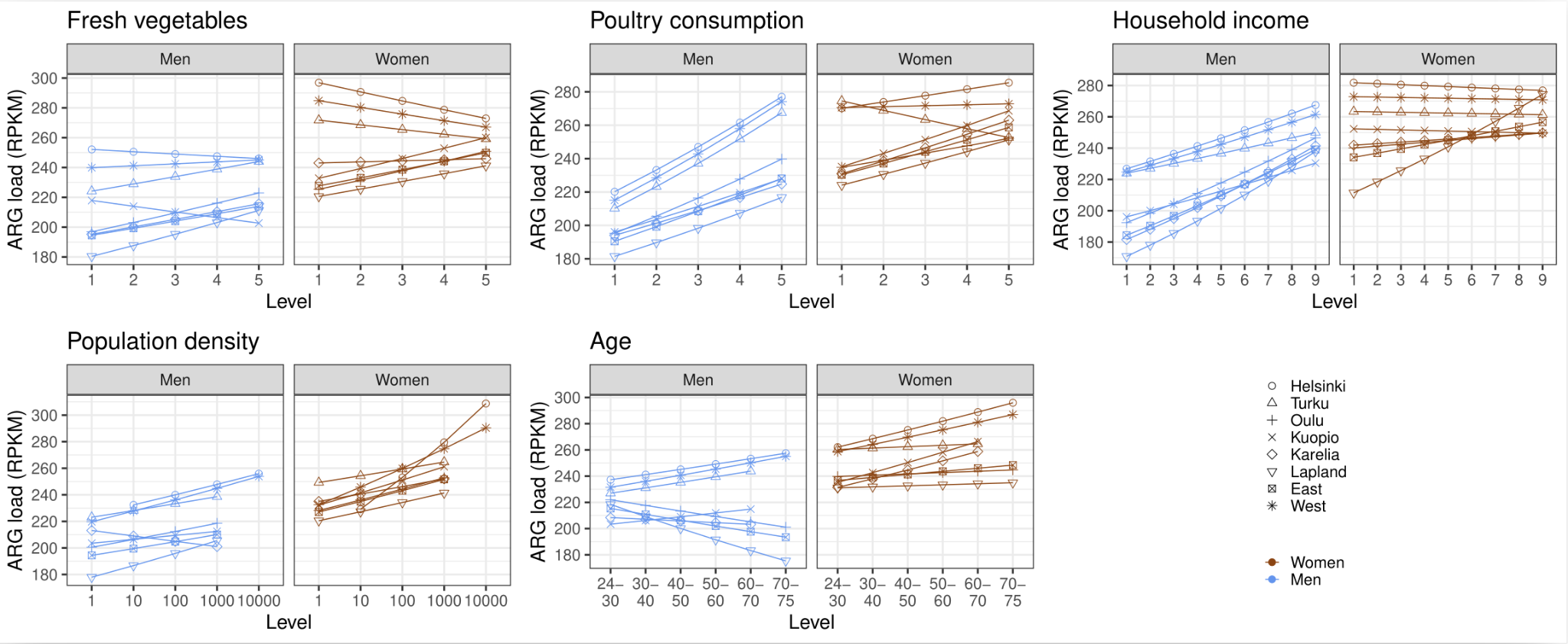

**Figure S2: Regional trends between ARG load and fresh vegetables and poultry consumption, household income, population density, and age group.** The figures show the linear model fit between the ARG load (log10 RPKM) and the indicated variable levels. Separate models for each region and gender were fitted using the R *lm* function. Western Finland covers the urban regions of Helsinki and Turku; Eastern Finland covers the other four regions. The effect sizes (slope) and significance estimates are shown in [Extended Data Table 4](https://docs.google.com/document/u/0/d/1bkZbyn2QTsXdjEgp8yTDV0qPeTUeNflWMRbBOTphw8Q/edit).

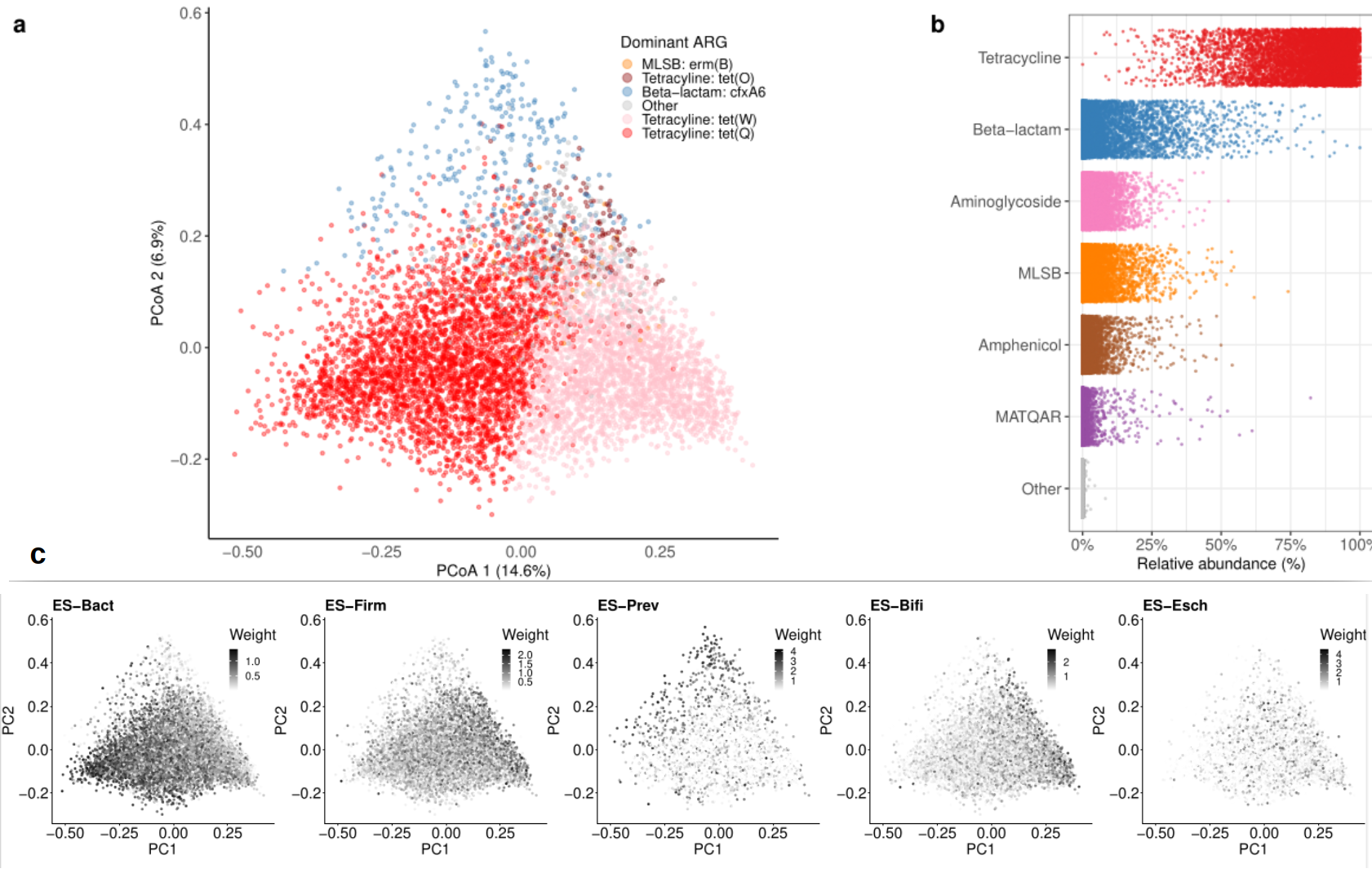

**Figure S3: Resistome landscape.** **a** Population landscape of human gut resistome composition (PcoA; Bray-Curtis index). The dominant (most abundant) antibiotic resistance gene for each sample is highlighted by color. **b** Relative abundance distribution for the most abundant ARG classes among the study population (N=7,095; see also Figure S4). **c** Resistome population variation landscape is linked to bacterial abundance variation. The ordination shows enterosignature weights across the resistome landscape (the normalized NMF score; see *Methods*). Each enterosignature corresponds to a particular set of co-abundant bacterial genera (Figure S6), and a higher weight indicates a stronger presence of the indicated enterosignature. See [Supplementary Table 1](https://docs.google.com/spreadsheets/d/1Vn-QHMeFmsuMh8yjpypN9UfioNf-xtgFhaJ-mN7Y2wk/edit" \l "gid=1822998243) for associations between the dominant bacterial families and ARG load.

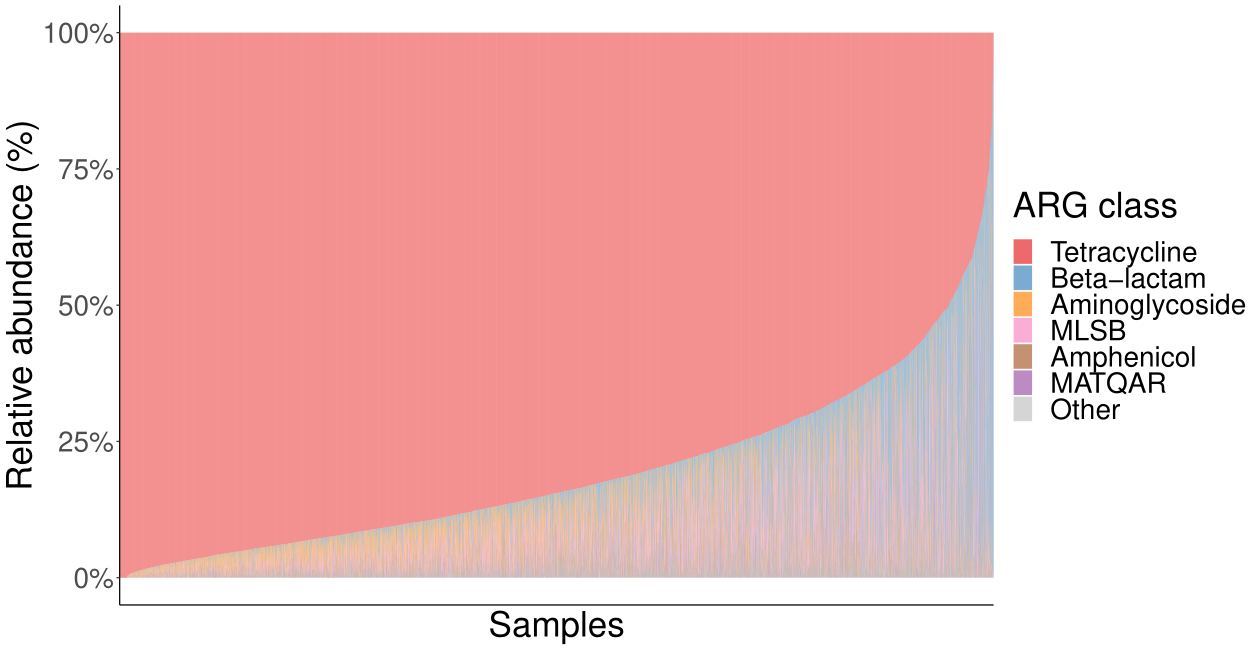

**Figure S4. Relative abundances of the antibiotic resistance gene classes in the FINRISK cohort (N=7,095).** The classes are defined by the antibiotic class for which the genes confer resistance to (Resfinder). Abbreviations: MLSB: “Macrolide, Lincosamide, Streptogramin B”; MATQAR: "Macrolide, Aminoglycoside, Tetracycline, Quinolone, Amphenicol, Rifamycin"

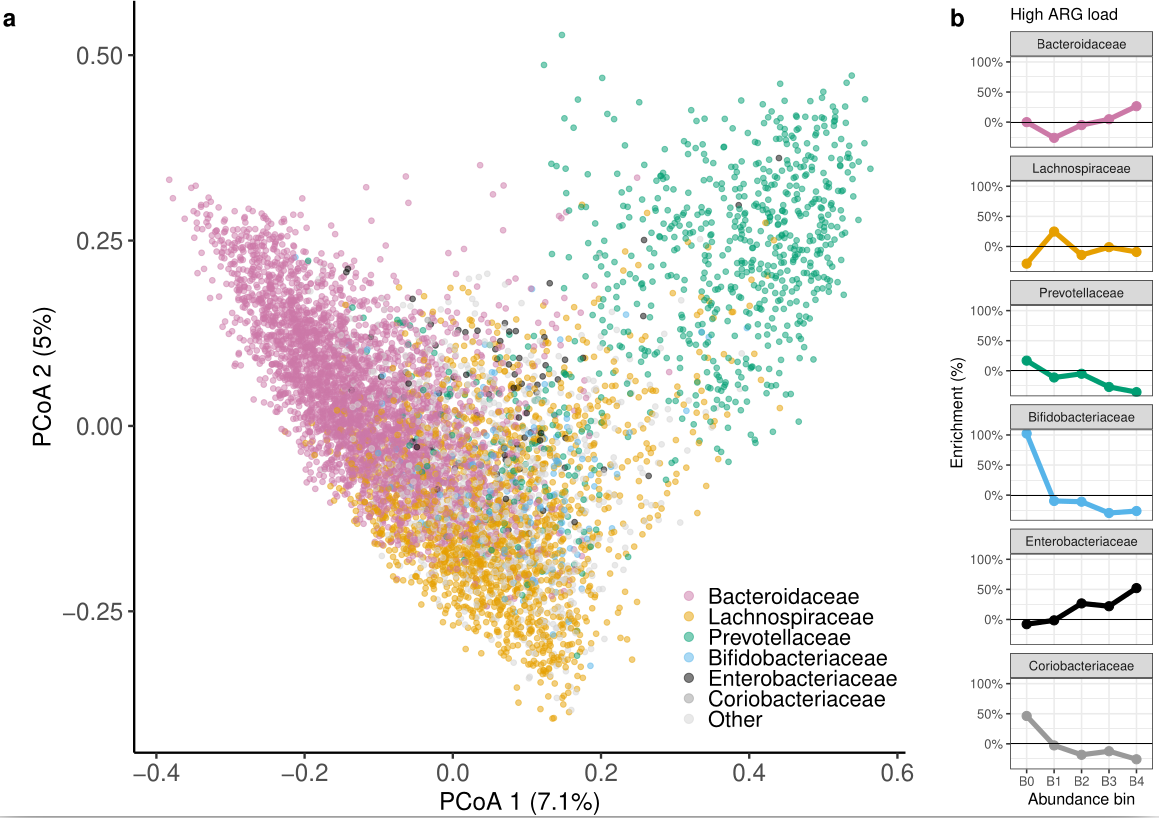

**Figure S5: Population landscape of microbial community composition and the observed ARG load. a** Population variation, or *landscape*, of the species-level microbiota composition among the 7,095 study participants (species-level PCoA; Bray-Curtis index). Each sample is colored by its most dominant bacterial family, and the families that are significantly associated with ARG load are highlighted ([Supplementary Table 1](https://docs.google.com/spreadsheets/d/1Vn-QHMeFmsuMh8yjpypN9UfioNf-xtgFhaJ-mN7Y2wk/edit#gid=1822998243); P<0.05); each sample is colored according to its dominant (most abundant) bacterial family. **b** The enrichment of individuals with high ARG load (top-10% quantile; >458 RPKM) across the abundance quantiles of each bacterial family (B0: not detected; B1-B4 25% abundance quartiles among individuals with detected signal).

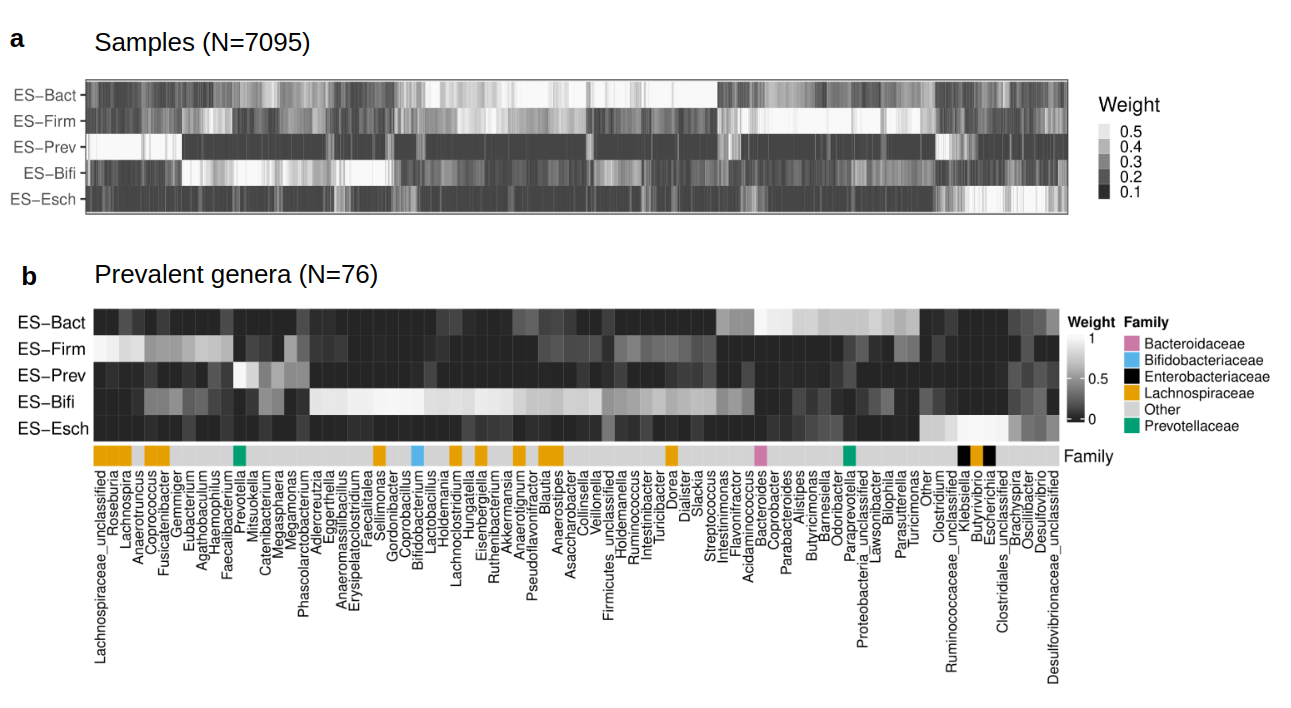
**Figure S6: Enterosignature profiles in FINRISK. a** Sample weights for each enterosignature (ES) across the 7095 FINRISK participants. The ES scores were normalized by the total sum score per sample. Values above the 90% quantile have been capped for the heatmap visualization. Each individual carries a unique mixture of enterosignatures. **b** Each of the five enterosignatures (ES) represents a mixture of prevalent genera; the heatmap indicates the relative weights for the prevalent genera in each ES (see [*Methods*](https://docs.google.com/document/d/1z36lt04Dct0uWIWn3eFdyGFqVy88wXPdm9fvUnW7hPQ/edit)); the bottom panel indicates their corresponding bacterial families (colors). Associations between each ES and species diversity, ARG diversity, and the total ARG load are provided in [Supplementary Table 2](https://docs.google.com/spreadsheets/d/1PgziatWXq1riYLGz0JMVUDU-GObuefs49ubsLtOTHSA/edit?usp=sharing). Associations between ES and the resistome: see Figure 4c (resistome characteristics).

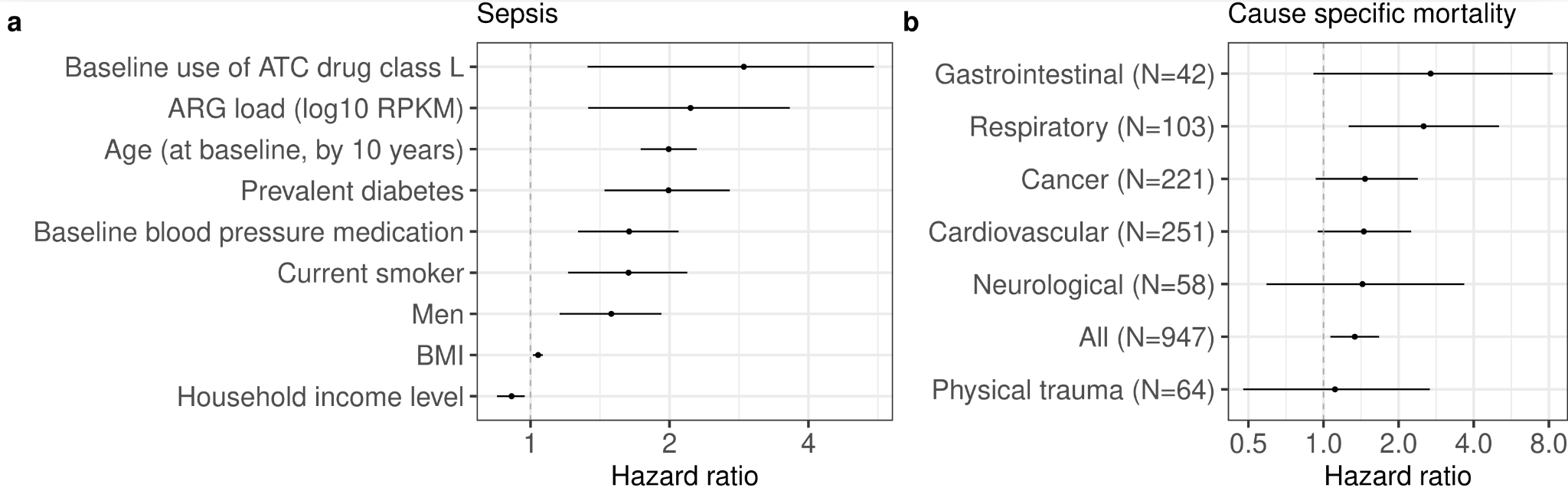

**Figure S7: Factors contributing to sepsis and cause-specific mortality events.**

**a** ARG load is associated independently with sepsis (probabilistic multivariate Cox proportional hazards; Extended Data Table 9). Variables whose 95% credible interval overlaps with 1 (no association) are excluded from the graph. **b** Associations between ARG load (log10 RPKM) and cause-specific mortality (Extended Data Table 8)**.** The median hazard ratio (HR) is shown for each variable, along with the 95% credible intervals. The models were adjusted for *Enterobacteriaceae* (*log10p* relative) abundance, age, smoking, gender, diabetes, use of antineoplastic and immunomodulating agents, body-mass index, self-reported antihypertensive medication, systolic blood pressure, prior antibiotics use (during six months before baseline), household income, and fresh salad and vegetable consumption (see [*Methods*](https://docs.google.com/document/d/1z36lt04Dct0uWIWn3eFdyGFqVy88wXPdm9fvUnW7hPQ/edit)).
